# Risk of Post-acute Symptoms and Conditions After SARS-CoV-2 Compared to Other Respiratory Viral Infections: A Systematic Review and Meta-Analysis

**DOI:** 10.64898/2026.04.11.26350682

**Authors:** Tomaz Fonseca Pinto, Andrea Santoro, Ana Laura Grossi de Oliveira, Thais Silva Tavares, André Almeida, Francesca Incardona, Giulia Marchetti, Alessandro Cozzi-Lepri, Jorge Pinto, Julia Fonseca de Morais Caporali

**Affiliations:** Programa de Pós-Graduação em Ciências da Saúde: Infectologia e Medicina Tropical, Faculdade de Medicina, Universidade Federal de Minas Gerais, Belo Horizonte, Minas Gerais, Brazil; Department of Health Sciences, Clinic of Infectious Diseases, San Paolo Hospital, ASST Santi Paolo E Carlo, University of Milan, Milan, Italy; Department of Public Health Education, Morehouse School of Medicine, Atlanta, GA, USA; Department of Internal Medicine 4, Unidade Local de Saúde São José, Centro Clínico Académico de Lisboa, Lisbon, Portugal; NOVA Medical School, Lisbon, Portugal; EuResist Network GEIE, Rome, Italy; InformaPRO S.R.L, Rome, Italy; Centre for Clinical Research, Epidemiology, Modelling and Evaluation (CREME), Institute for Global Health, UCL, London WC1E 6BT, UK; Department of Pediatrics, Universidade Federal de Minas Gerais, Belo Horizonte, Minas Gerais, Brazil

## Abstract

**Background:** How post-COVID-19 condition (PCC) differs from post-acute infection syndromes (PAIS) caused by other respiratory viruses remains uncertain. Comparing these conditions may clarify whether post-acute symptoms reflect specific consequences of SARS-CoV-2 infection or broader post-viral mechanisms.

**Methods:** We conducted a systematic review and meta-analysis of cohort studies comparing persistent symptoms or conditions in adults after SARS-CoV-2 infection with those following other acute respiratory viral infections. PubMed, Embase, and Scopus were searched. Random-effects models were used to estimate pooled risks.

**Results:** Among 9,371 records screened, 22 studies were included and 14 contributed to the meta-analysis. Increased risk after SARS-CoV-2 infection was observed for pulmonary embolism, abnormal breathing, fatigue, hemorrhagic stroke, memory loss/brain fog, and palpitations; heart rate abnormalities showed borderline significance. For most other outcomes pooled estimates were inconclusive.

**Conclusions:** Only a subset of outcomes appears more frequent after SARS-CoV-2 infection, suggesting many symptoms attributed to PCC may reflect broader post-viral syndromes.

## Introduction

Post-acute symptoms and conditions following SARS-CoV-2 infection were recognized during the first year of the COVID-19 pandemic as a significant health problem affecting patients recovering from severe, moderate, or mild disease. Early studies, however, showed substantial heterogeneity in the definitions of symptoms, conditions, onset, and duration of post-acute outcomes. To standardize these criteria, the World Health Organization developed a Delphi consensus in 2021 defining post-COVID-19 condition (PCC) as symptoms occurring in individuals with probable or confirmed infection, usually three months after onset, lasting at least two months, and not explained by an alternative diagnosis. Common manifestations include fatigue, shortness of breath, and cognitive dysfunction, which may persist from the acute illness or appear after recovery and may fluctuate over time.^1^

Although numerous studies and systematic reviews have investigated PCC, important uncertainties remain. Its definition varies across organizations,^1–3^ and terminology is inconsistent, with PCC also referred to as Long COVID, post-acute COVID-19 syndrome (PACS),^4^ and post-acute sequelae of COVID-19 (PASC).^5^ Reported incidence ranges from 4% to 85%,^6–8^ depending on the population studied. The most frequently reported symptoms include fatigue, dyspnea, cognitive dysfunction or brain fog, sleep disturbances, cough, tachycardia, pain, loss of smell or taste, depression, anxiety, and fever.^1^ In addition, cardiovascular, neurological, psychiatric, gastrointestinal, and metabolic disorders have been described as sequelae.^5^ The pathophysiology remains unclear, with proposed mechanisms including immune dysregulation, viral persistence or reactivation of other viruses, gut dysbiosis, inflammation, autoimmunity, coagulation abnormalities, and dysfunctional neurological signalling.^6^

Uncertainty surrounding PCC is widely recognised in the literature, ^6,9,10^ and partly reflects substantial methodological and outcome heterogeneity across studies. Common limitations include selection bias in case–control designs and the absence of appropriate control groups.^11,12^

A recent systematic review on PCC tried to reduce recall and selection biases by focusing on prospective studies.^13^ They found that more than half of COVID-19 survivors experienced PCC during at least 26 weeks of follow-up. Another meta-analysis including more than 14 million participants found an increased risk for 39 of 40 symptoms when comparing SARS-CoV-2–infected individuals with asymptomatic uninfected controls.^14^

However, the review excluded symptomatic control patients with other respiratory infections. This represents a missed opportunity, as comparisons between PCC and post-acute infection syndrome (PAIS) from other respiratory viruses could provide valuable insights.

Even though PAIS has been far less studied than PCC, it was recognized long before the COVID-19 pandemic as chronic sequelae following acute infections caused by viruses such as influenza, SARS, Epstein–Barr virus, varicella-zoster virus, Ebola virus, and dengue virus, as well as by non-viral pathogens. A recent review of PAIS in adults concluded that, despite different infectious triggers, these syndromes share several symptoms—including fatigue, myalgia, irritability, depression, neurocognitive and sensory impairment, and flu-like symptoms—many of which overlap with PCC. Some manifestations, however, may be pathogen-specific, possibly reflecting distinct tropisms and mechanisms, as may occur with SARS-CoV-2. Proposed, non-mutually exclusive mechanisms for PAIS resemble those suggested for PCC and include pathogen persistence or remnants, autoimmunity, gut dysbiosis, pathogen reactivation, and impaired repair of acute tissue damage in organs such as the heart, lungs, brain, and endothelium.^15^

Comparing PAIS with PCC may clarify whether post-acute symptoms and conditions reflect unique consequences of SARS-CoV-2 infection or shared pathogenic pathways.^12^ This distinction is important for advancing research on pathophysiology, biomarkers, and treatments, while also increasing recognition of the broader category of PAIS.^6^

This systematic review and meta-analysis compared the frequency of post-acute symptoms and conditions in adults after SARS-CoV-2 infection with those following other acute respiratory viral infections.

## Methods

### Search Strategy and Information Sources

We conducted a systematic review and meta-analysis comparing the incidence of post-acute outcomes after SARS-CoV-2 infection with those following other respiratory viruses in adults and older individuals. The protocol followed PRISMA guidelines (appendix pp 2–5) and was registered in PROSPERO (CRD42024560894).^16^

From June 21, 2024, to April 13, 2025, we searched MEDLINE (PubMed), Embase, and Scopus without date restrictions using Medical Subject Headings (MeSH), Emtree terms, and DeCS descriptors combined with Boolean operators (appendix pp 6–7). Searches were not restricted by language. Search alerts were activated to capture newly published studies, and reference lists of retrieved articles and reviews were screened manually.

### Eligibility criteria and study selection

Eligible studies were peer-reviewed observational studies or randomized controlled trials comparing adults (≥18 years) with SARS-CoV-2 infection to comparator groups with viral acute respiratory infection (viral ARI) or influenza-like illness (ILI), including infections caused by influenza A or B, adenovirus, metapneumovirus, parainfluenza viruses, respiratory syncytial virus, or rhinovirus. Studies focusing exclusively on children, pregnant women, or immunocompromised populations were excluded.

Outcomes of interest were post-acute symptoms or conditions compatible with post-COVID-19 condition (PCC). To ensure comparability, we used the core outcome set established by the International Delphi consensus for PCC (appendix pp 8–9),^17^ which was also applied to post-acute infection syndromes (PAIS) in the absence of a comparable consensus definition. Eligible studies reported the frequency of at least one outcome from this core set with ≥3 months of follow-up after acute infection.

Two reviewers independently screened titles and abstracts (TFP and ALGO) and full texts (TFP and AS), with disagreements resolved by consensus with a third author (JFMC). Studies with incomplete quantitative data or without peer review were excluded. Screening was performed using Rayyan AI™ after removal of duplicates.

### Data extraction

Data were extracted using a standardised Microsoft Excel® form. Two investigators (TFP and AA) independently collected study characteristics, including author, year, country, study design, setting, sample size, age, sex, and follow-up duration. They also extracted post-acute outcomes (symptoms and conditions), which were harmonised and coded according to the Delphi consensus classification (appendix pp 12–16). For studies included in the meta-analysis, outcome-level data—including cohort sizes and outcome frequencies for COVID-19 and comparator groups—were extracted and used to compute risk ratios for quantitative synthesis (appendix pp 17–27).

### Quality assessment

Two investigators (TFP and ALGO) independently assessed study quality using the Newcastle–Ottawa Scale (NOS), which evaluates selection (maximum four stars), comparability (two stars), and outcome assessment (three stars) (appendix pp 10–11). Studies scoring ≥7 stars were considered high quality. Disagreements were resolved by consensus or consultation with a third reviewer (JFMC).

### Synthesis and Data Analysis

Outcomes were eligible for meta-analysis when at least three studies reported sufficiently comparable data.

Meta-analyses were conducted separately for each outcome using risk ratios (RRs) comparing COVID-19 with other viral ARI or ILI. Random-effects models with Hartung–Knapp adjustment were applied using the meta and metafor packages in R (version 4.3).^19,20^ Pooled estimates were presented as log RRs with 95% CIs in forest plots.

Heterogeneity was assessed using Cochran’s Q test and the I² statistic.^21^ Between-study variance was estimated with τ². Publication bias was assessed only for outcomes with ≥10 studies using funnel plots, Egger’s regression, Begg’s test, and the trim-and-fill method.^22–25^ Additional exploratory analyses included meta-regression and influence diagnostics to evaluate heterogeneity and robustness.^26^

## Results

### Characteristics of the included studies

A total of 9,371 records were identified, of which 249 were assessed in full text after screening. Twenty-two studies met inclusion criteria for the systematic review, and 14 provided sufficient data for meta-analysis (figure 1).^27–48^

**Figure 1.**
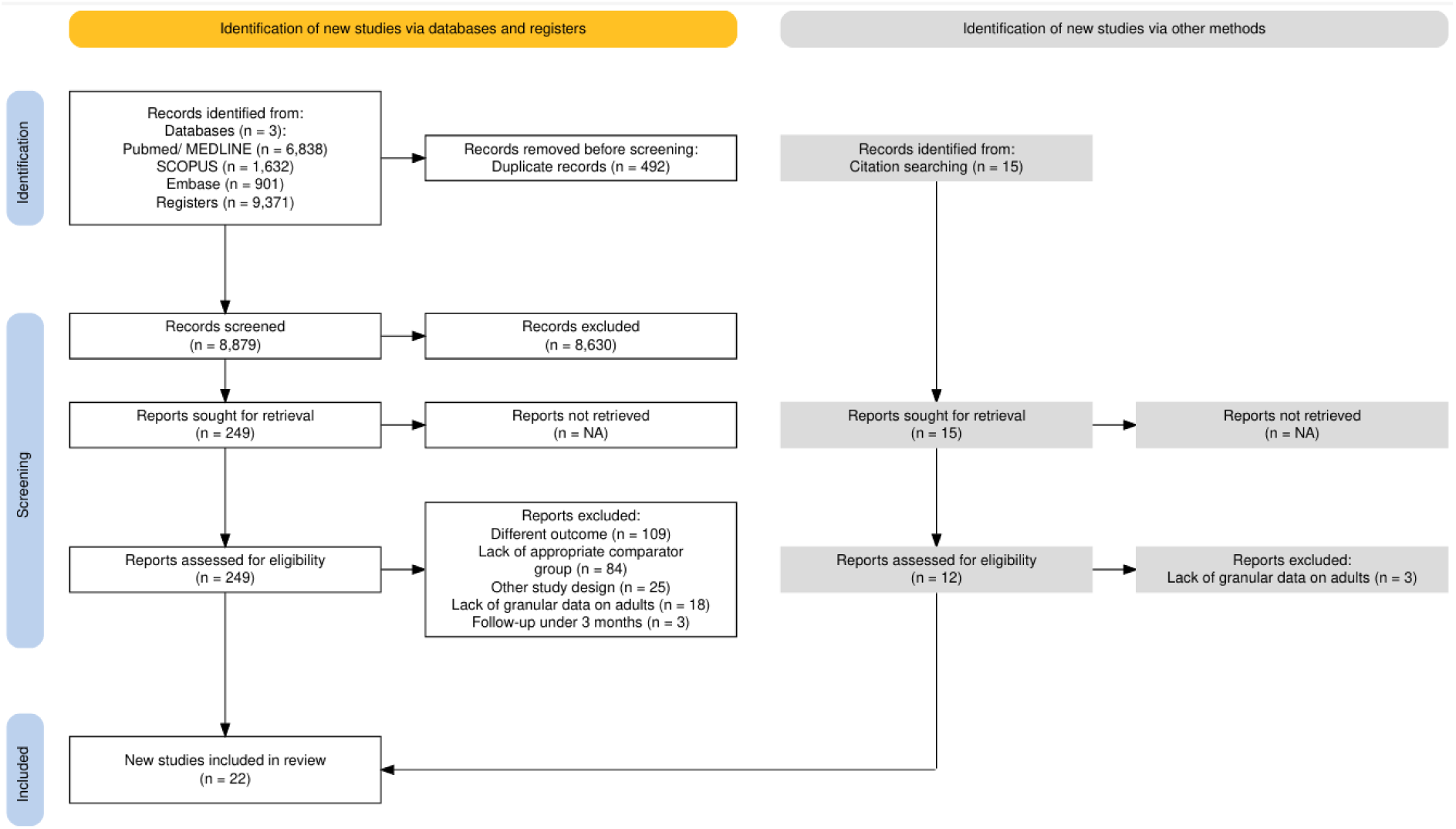
PRISMA Flow chart. **Alt text:** PRISMA flow diagram showing the study selection process. From 9,371 records identified in PubMed/MEDLINE, Scopus, and Embase, 492 duplicates were removed. Of 8,879 screened records, 249 full texts were assessed for eligibility. Twenty-two studies met the inclusion criteria for the systematic review after exclusions due to different outcomes, inappropriate comparator groups, study design, insufficient adult data, or follow-up under three months.

All included studies were multicentre retrospective cohorts conducted in North America (10), Asia (4), Europe (3), or across multiple continents (5), several using large health-data networks such as TriNetX. Sample sizes ranged from fewer than 10,000 to more than two million participants per group. Comparator groups most commonly involved ILI (15 studies), followed by influenza (6) and RSV (1). Most cohorts included both inpatient and outpatient populations (14), with fewer restricted to inpatients (6) or outpatients (2) (table 1).

**Table 1.** Baseline characteristics of included studies.

| | Authors/year/<br>Country | Continent | Setting | Sample size<br>(COVID/<br>control) | Age<br>(mean ± SD) | Female sex<br>(%) | Follow-up<br>Duration<br>(months) | Quality<br>assessment<br>Assessment<br>tool:<br>Newcastle-Ot<br>tawa Scale | Matching<br>strategies |
| --- | --- | --- | --- | --- | --- | --- | --- | --- | --- |
| 1 | Liu, Ting-Hui et al.<br>December, 2023.<br>Tainan, Taiwan | Africa, Asia, Europe,<br>Middle East, North<br>America, South<br>America | Inpatient | 6614/6614<br>(COVID/ Flu-<br>contemporary) | 62.7 (± 16.7)/<br>63.1 (±17.9) | 47.55/48.1 | 3-6 | 9 | 1:1 propensity<br>score (PS):<br>demographic,<br>socioeconomic,<br>comorbidities |
| 2 | Xie Y, Choi T, Al-Aly<br>Z. December,2023.<br>Saint Louis, USA | North America | Inpatient | 81280/ 10985<br>(COVID/ILI) | 70.80 (±12.7)/<br>70.53 (±12.7) | 5.1/5.1 | 18 | 7 | PS inverse<br>probability:<br>demographic,<br>socioeconomic,<br>comorbidities, prior<br>healthcare<br>utilization,<br>laboratory tests,<br>vaccination status,<br>vital measurements |
| 3 | Velásquez EE et al.<br>July, 2024. Palo Alto,<br>USA | North America | Outpatient | 27960/27960<br>(COVID/ILI) | 55.68 (±17.10)/<br>55.54 (±17.09) | 56.7/56.8 | 6 | 9 | 1:1 PS:<br>demographic,<br>socioeconomic,<br>prior healthcare<br>utilization,<br>comorbidities |
| 4 | Han, M et al. May<br>2024. Seoul, Korea. | Asia | Both | 1656/1656<br>(COVID/ILI) | 37.98 (± 13.79)/<br>57.17 (±19.39) | 48.2/ 54.2 | 3 | 8 | 1:1 PS:<br>demographic,<br>comorbidities |
| 5 | Whittaker H R et al.<br>Nov 2021. London<br>UK | Europe | Both | 437943<br>(COVID<br>outpatient)/<br>18059 (COVID<br>inpatient)/2180<br>3 (ILI<br>outpatient) | 42.91(± 17.10)/<br>47.12 (±15.60) | 55.6 (COVID<br>outpatient)/<br>49.4 (COVID<br>inpatient)/ 60.1<br>(ILI) | 3-9 | 8 | Stratification by<br>age, gender,<br>comorbidities and<br>risk factors. Not<br>matched |
| 6 | Quinn K L et al. June, 2023. Toronto, Canada | North America | Inpatient | 26499/17516 (COVID/ILI) | Median age (IQR): COVID-69 (55 - 81); ILI- 69 (55-81) | 46.4; 52.3 | 12 | 9 | PS-based: demographic, socioeconomic, comorbidities, severity of illness, prior healthcare utilization, Frailty score, vaccination status |
| 7 | Tesch F et al. October, 2024. Dresden, Germany | Europe | Both | 573791/569154 (COVID/Flu) | 50.78 (± 20.5)/ 48.76 (± 18.2) | 58.3/ 58.3 (COVID/Flu) | 3-15 | 9 | 1:1 PS matched: demographic, socioeconomic, severity of illness, comorbidities |
| 8 | Wee L E et al. Dec, 2023. Singapore, Singapore | Asia | Both | 104179/666575 (COVID delta/ILI); 375903/ 619379 (COVID omicron/ILI)--contemporary | 49 (± 16.90) Delta/ 49 (±17.46) ILI delta period; 48 (±17.56) Omicron/ 48 (±17.41) ILI Omicron period | 45.6 (Delta)/ 51.7 (ILI Delta period)/ 51.8 (Omicron)/ 51.9 (ILI Omicron period) | 10 | 9 | PS matched: demographic, socioeconomic and comorbidities |
| 9 | Havenon A et al. April,2024. New Haven, USA | North America, Europe, Asia | Inpatient | 77272/77272 (COVID/Flu) | 51 (±19.7)/ 51 (±19.7) | 57.7/ 57.7 | 12 | 8 | 1:1 PS-matched: demographic, socioeconomic, comorbidities |
| 10 | Zarifkar P et al. June,2022. Copenhagen, Denmark | Europe | Both | 8013/4142 (COVID inpatient/Flu inpatient); 35362 / 3960 (COVID outpatient/Flu outpatient) | 66 (COVID In)/68 (Flu in); 48y (out); 52y (out) | 44.5/ 54.0; 59.1/ 59.9 | 12 | 8 | stratification by age, gender, comorbidities and risk factors. Not matched |
| 11 | Liu TH et al. November, 2023. Tainan, Taiwan | Africa, Asia, Europe, Middle East, North America, South America | Both | 9220/9220 (COVID/ILI)--contemporary | 71.2 (±7.0)/ 70.5 (±8.0) | 56.5/52.51 | 3-6 | 8 | 1:1 PS matching: demographic, socioeconomic, comorbidities |
| 12 | Al-Aly Z et al. April,2021. Saint Louis, USA | North America | Inpatient | 13654/13997 (COVID/ILI) | 69.20 (±12.82)/ 69.20 (±12.97) | 5.81/5.64 | 6 | 8 | PS-based weighted: predefined and high-dimensional covariates |
| 13 | Baskett W et al. December, 2022. Columbia, USA | North America | Both | 17487/17487 (COVID/ILI)--contemporary | 50.25 (±18.7)/ 50.84(± 18.5) | 66.1/66.1 | 12 | 8 | PS-based: demographics and comorbidities, severity of illness |
| 14 | Cohen K et al. December, 2021. Minnetonka, USA | North America | Both | 73490/73490 (COVID/ILI) | 65-74: 42.99/ 42.99<br>>74: 55.36/ 55.36 | 56.28/56.28 | 3w-5m | 8 | 1:1 PS matched: demographic, socioeconomic, comorbidities, prior healthcare utilization |
| 15 | Cabrera C I et al. November, 2023. Cleveland, USA | Africa, Asia, Europe, Middle East, North America, South America | Both | 2544070/ 101886 (COVID/ILI) | 46.1 (± 17.4)/ 42.2 (±18.8) | 56.2/59.6 | 12 | 8 | PS matched: demographics |
| 16 | Donnachie E et al. September 2022. Munchen, Germany | Europe | Outpatient | 348087/55641 (COVID/ILI) | 46.11 (±16.7)/ 45.25 (±16.2) | 54.1/54.1 | 24 | 9 | PS-based weighting: demographics, comorbidities |
| 17 | Daugherty S E et al. April, 2021. Minneapolis, USA | North America | Both | 181613/181613 (COVID/Flu) | 41.9 (13.9)/ 42.8 (13.6) | 53.1/ 52.9 | 108d (IQR 66-145d)/ 108d (IQR 65-141d) | 8 | PS matched: demographic, socioeconomic, comorbidities, prior healthcare utilization |
| 18 | Xie Y et al. January, 2022. Saint Louis, USA | North America | Both | 132852/60283 (COVID outpatient/Flu outpatient); 20996/ 11924 (COVID inpatient/Flu inpatient) | 63.06 (± 16.18)/ N/A<br>Flu group data non available. | 10.78/ N/A | 10-20 | 7 | PS-based weighted: predefined covariates and high-dimensional covariates |
| 19 | Fung KW et al. April, 2023. Bethesda, USA | North America | Both | 234688/ 123736 (COVID outpatient/ Flu outpatient); 58484/ 16,961( COVID inpatient/ Flu inpatient) | Median age (IQR)- Outpatient: COVID-75.0 (70-82); Flu- 74.0 (70-81)// Inpatient: COVID- 78.0 (72-85); 81.0 (74-88) | Outpatient: COVID- 57.7/ Flu- 60.1 Inpatient: COVID 50.5/ Flu 58.2 | 1-3 | 8 | Generalized linear model with logit link adjusting: demographics, socioeconomic, comorbidities, prior healthcare utilization |
| 20 | Ward A et al. January, 2022. California, USA | North America | Both | 417975/345934 (COVID/ILI) | Median age (IQR): 57 (40-72)/ 47 (32-61) | 61.0/ 66.0 | 3 | 8 | PS matched: demographic, severity of illness, comorbidities, risk factors, medication use |
| 21 | Wee E L et al. April, 2025. Singapore, Singapore | Asia | Inpatient | 70628/ 12007 (COVID/ Flu and RSV) | 59.71 ($\pm 16.11$) / 56.64 ($\pm 17.41$) | 51.0 /54.7/ 60.5 | 1-10 | 9 | PS-based overlap weighted and regression adjusted: demographic and socioeconomic status, severity of illness, prior healthcare utilization, comorbidities. |
| 22 | Lee, H et al. Feb 2022. Seoul, South Korea | Asia | Both | 18778/605010 (COVID/ILI) | 50.33 ($\pm 17.20$)/ 47.90 ($\pm 16.83$) | N/A<br>Data not available | COVID: 7<br>ILI: 12 | 8 | Stratification by age, gender, comorbidities and risk factors. Not matched |

Age distributions were heterogeneous but similar between groups, with most COVID-19 cohorts including middle-aged (10) or older adults (8). Female participation was generally balanced (30–60%). Follow-up ranged from <6 months (8 studies) to >1 year (4), with 10 reporting 6–12 months. Overall methodological quality was high: 20 studies scored 8–9 on the NOS, with a mean score of 8.14 (table 1). Most studies (15) reported both post-acute symptoms and conditions, while five reported only conditions and two only symptoms (table 1).

### Occurrence of post-acute symptoms after COVID-19

Post-acute symptoms after COVID-19 varied across studies, with cognitive impairment, fatigue, and respiratory symptoms most consistently elevated. Abnormal breathing was reported in nine of 17 studies and showed the highest relative risks in some cohorts. Chest or throat pain was reported in eight studies with modest increases. Cognitive impairment or memory loss (15 studies) and fatigue or malaise (15 studies) were consistently elevated. Headache or migraine (14 studies) and myalgia or arthralgia (10 studies) showed heterogeneous associations. Palpitations were less consistently reported (5 studies), whereas sensory disturbances including anosmia or dysgeusia appeared in eight studies, occasionally with high relative risks. Sleep disorders (10 studies) and visual or auditory disturbances (5 studies) showed variable findings across cohorts (table 2).

**Table 2.** Occurrence of selected post-acute symptoms in COVID-19 and comparison groups, by study.

|  | Study | Estimates | Comparison Groups Description | Post-acute symptoms |  |  |  |  |  |  |  |  |  |  |
| --- | --- | --- | --- | --- | --- | --- | --- | --- | --- | --- | --- | --- | --- | --- |
|  |  |  |  | Abnormal breathing (9/17) | Chest/throat pain (8/17) | Memory loss/brain fog (15/17) | Cough (7/17) | Fatigue/malaise (15/17) | Headache/migraine (14/17) | Myalgia / arthralgia (10/17) | Palpitations (5/17) | Sensory disorders (anosmia/dysgeusia- 8/17) | Sleep disorders (10/17) | Visual/Auditory Disturbance (5/17) |
| 1 | Liu, Ting-Hui et al. December, 2023. Tainan, Taiwan | Cumulative incidence (%); HR (95% CI; p value) | Flu (contemporary cohort) | 6.40/4.27; HR 1.51 (1.25-1.82; <0.01) | 2.97/2.43; HR 1.23 (0.94-1.60; 0.13) | 1.74/0.96; HR 1.82 (1.24-2.67; <0.01) | 2.03/1.87; HR 1.07 (0.78-1.48; 0.65) | 3.75/2.50; HR 1.49 (1.16-1.91; 0.02) | 1.12/1.09; HR 1.00(0.66-1.52; 0.99) | 0.5/0.36; HR 1.40 (0.71-2.75; 0.33) | 0.65/0.56; HR 1.14 (0.65-2.01; 0.64) | NA | 1.35/1.03; HR 1.29 (0.86-1.94; 0.21) | NA |
| 2 | Xie Y, Choi T, Al-Aly Z. December,2023. Saint Louis, USA | Incident rate per 100 person at 540 days; HR (95% CI). | ILI (historical cohort) | 31.77/33.68; HR 0.93 (0.90 - 0.97) | NA | 10.07/8.11; HR 1.25 (1.17-1.35 ) | 14.59/24.45; HR 0.56 (0.54-0.59) | 35.61/ 31.21; HR 1.18 (1.13-1.22) | Headache: 1.77/ 1.46; HR 1.22 (1.02- 1.45) Migraine: 2.73/ 2.23; HR 1.23 (1.06- 1.41) | Arthralgia: 22.61/ 21.58; HR 1.05 (1.01-1.1) Myalgia : 23.82/ 14.31; HR 1.76 (1.67- 1.86) | NA | Anosmia: 0.23/ 0.07; HR 3.13 (1.47- 6.67) Dysgeusia: 0.11/ 0.08; 1.34 (0.65- 2.78) | 25.49/ 21.79; HR 1.2 (1.15- 1.25) | Loss of hearing: 15.53/ 15.19; 1.02 (0.97- 1.08) Visual disturbance: 9.2/ 8.13; 1.14 (1.05- 1.23) |
| 3 | Velásquez EE et al. July, 2024. Palo Alto, USA | Cumulative incidence (%); Effect size | ILI (historical cohort) | 4.4/3.2; Effect size 0.03 | NA | 2.1/ 2.2; Effect size 0.02 | NA | 3.7/ 3.2; Effect size 0.00 | 2.3/ 1.7; Effect size 0.02 | NA | NA | NA | 3.6/ 3.2; Effect size 0.02 | 0.3/ 0.4; Effect size 0.02 |
| 4 | Han, M et al. May 2024. Seoul, Korea. | Relative risk (95% CI, p value) | ILI (contemporary cohort) | NA | 0.63 (0.37-1.05; 0.08) | 0.33 (0.14-0.71; 0.01) | 1.00 (0.51-1.97; 1.00) | 0.57 (0.15-1.89; 0.39) | 0.50 (0.34-0.73; <0.01) | Arthralgia: 0.77 (0.48-1.20; 0.25) | NA | NA | 2.50 (1.02-7.01; 0.06) | Ocular symptoms: 1.35 (0.96-1.91 ; 0.09) |
|  |  |  |  |  |  |  |  |  |  | Myalgia : 1.03 (0.72-1.49; 0.85) |  |  |  |  |
| 5 | Whittaker H R et al. Nov 2021. London UK | Adjusted Hazard ratios (95% CI) for difference in event rates after index date compared with the same patient 12 months before | COVID outpatient/ COVID inpatient/ ILI outpatient (historical cohort) | 1.39 (1.31-1.47)/ 1.88 (1.62-2.19)/ 0.64 (0.35-1.19) | 1.22 (1.14-1.3)/ 1.39 (1.11-1.75)/ 1.06 (0.91 - 1.25) | 1.15 (0.9-1.46)/ 1.66 (1.00-2.77)/ 1.17 (1.03-1.33) | 0.49 (0.46-0.51)/ 0.67 (0.57-0.78)/ 1.15 (0.87-1.52) | 1.64 (1.53-1.76)/ 2.52 (1.81- 3.51)/ 0.89 (0.69-1.15) | 1.08 (1.02-1.14)/ 0.83 (0.62-1.12)/ 0.81 (0.69- 0.94) | Arthralgia: 0.93 (0.91-0.96)/ 0.85 (0.75-0.97)/ 0.96 (0.90-1.04)<br>Myalgia : 1.89 (1.63-2.20)/ 2.07 (1.16-3.69)/ 1.1 (0.76-1.6) | 1.42 (1.27-1.59)/ 2.55 (1.61-4.05)/ 1.37 (0.67-2.82) | 5.28 (3.89-7.17)/ 2.02 (0.50-8.08)/ 0.95 (0.8-1.12) | 1.50 (1.33-1.69)/ 2.17 (1.36-3.49)/ 1.43 (0.88-2.33) | Tinnitus: 1.36 (1.14-1.62)/ 1.43 (0.58-3.54)/ 1.14 (0.56-2.35) |
| 6 | Tesch F et al. October, 2024. Dresden, Germany | Cumulative incidence (%); Incidence rate ratio 12-15m after index date | Flu (historical cohort) | 2.17/ 0.85 IRR: 2.04 (1.90-2.19) | 0.04/ 0.01 IRR: 2.64 (1.55-4.50) | 0.46/ 0.39 IRR: 1.19 (1.06-1.33) | NA | Malaise/exhaustion: 2.15/ 1.35; IRR 1.63 (1.54-1.73)<br>Chronic Fatigue Syndrome: 0.37/ 0.05; IRR 3.50 (2.92-4.19) | NA | NA | NA | NA | NA | NA |
| 7 | Wee L E et al. Dec, 2023. Singapore, Singapore | Cumulative incidence (%); Adjusted HR (95 CI; p value) | ILI (contemporary- Delta period and Omicron period) | NA | NA | Delta: 0.22/ 0.12; HR 1.63(1.39-1.92; <0.01)<br>Omicron : 0.18/ 0.13; HR 1.24(1.12-1.38; <0.01) | NA | Delta: 0.22/ 0.12; HR 1.63(1.39-1.92; <0.01)<br>Omicron: 0.18/ 0.13; HR 1.24(1.12-1.38; <0.01) | Delta: 0.03/ 0.03; HR 0.81 (0.53-1.24)<br>Omicron: 0.03/ 0.03; HR 1.10 (0.86-1.39) | NA | NA | NA | Delta: 0.11/ 0.13; HR 0.84 (0.68-1.04)<br>Omicron: 0.12/ 0.13; HR 0.98 (0.87-1.10) | Hearing abnormalities: - Delta: 0.11/ 0.13; HR 0.90 (0.73-1.11; NA)<br>- Omicron: 0.15/0.14; HR 1.02 (0.92-1.14 ; NA) |
|  |  |  |  |  |  |  |  |  |  |  |  |  |  | Vision abnormalities<br>- Delta: 0.05/ 0.04; HR 1.05 (0.77-1.43)<br>- Omicron: 0.05/ 0.03; HR 1.11 (0.93-1.32; NA) |
| 8 | Havenon A et al. April, 2024. New Haven, USA | Cumulative incidence (%); HR (95% CI; p value) | Flu (historical cohort) | NA | NA | NA | NA | NA | 1.97/ 3.16; HR 0.65 (0.60–0.69; <0.01) | NA | NA | NA | NA | NA |
| 9 | Liu TH et al. November. 2023. Tainan, Taiwan | Cumulative incidence (%); HR (95 CI) | ILI (contemporary cohort) | 7.16/ 3.56; HR 2.05 (1.76-2.40) | 2.97 / 2.41; HR 1.23 (0.99-1.53) | 2.43/ 1.47; HR 1.67 (1.30-2.15) | 2.94/ 2.38; HR 1.25 (1.01-1.55) | 4.6/ 2.93; HR 1.587 (1.322-1.905) | 1.49/ 1.39; HR 1.06 (0.79- 1.42) | 0.62/ 0.75; HR 0.82 (0.53-1.28) | 0.98/ 0.90; HR 1.09 (0.76-1.57) | NA | 1 / 0.75; HR 1.34 (0.92-1.95) | NA |
| 10 | Al-Aly Z et al. April, 2021. Saint Louis, USA | Incidence rate per 1000 person-years ; HR (95% CI; p value) | ILI (historical cohort) | NA | 57.98/ 58.39; HR 0.99 (0.87-1.13, 0.91) | 54.69 / 38.53; HR 1.43 (1.24-1.65; 0.50) | NA | 123.85/ 87.36; HR 1.45 (1.31-1.59; 1.0) | 25.68/ 24.03; 1.07 (0.89- 1.29, >0.1) | NA | NA | NA | 126.02/ 110.76; HR 1.15 (1.03-1.27, < 0.01) | 43.67/ 55.13; HR 0.79 (0.68-0.91, < 0.01) |
| 11 | Baskett W et al. December, 2022. Columbia, USA | Cumulative incidence (%); OR (95% CI; p value) | ILI (contemporary cohort) | 15.4/ 13.6; OR 1.19 (1.11–1.27; <0.01) | 13.7/ 12.7; OR 1.09 (1.02–1.16; 0.01) | 7.4 / 6.6; OR 1.13 (1.03–1.24; 0.01) | 13.3/ 14.8; OR 0.87 (0.82–0.93; <0.01) | 12.5/ 11.5; OR 1.09 (1.02–1.17; <0.01) | 14.6/ 14.8; OR 0.97 (0.91–1.04; <0.5) | 23.36/ 21.63; OR 1.13 (1.07–1.19; <0.01) | 4.6/ 3.4; OR 1.42 (1.27–1.60; <0.01) | 0.3/ 0.3 OR 0.85 (0.58–1.24; <0.5) | 6.7/ 6.8; OR 0.97 (0.88–1.07; 0.50) | 0.90/ 0.70; OR 1.18 (0.93–1.51; 0.18) |
| 12 | Cohen K et al. December, 2021. Minnetonka, USA | Incident rates per person-years ; HR (95% CI; p value) | ILI (historical cohort) | NA | NA | 0.1715/ 0.1623; HR 1.03 (0.94-1.14; 1) | NA | 0.3365/ 0.3251; HR 1.02 (0.94- 1.1; 1) | 0.0095/ 0.0111; HR 0.86 (0.61-1.21; 1) | 0.2401/ 0.2681; HR 0.88 (0.8-0.96; 1) | NA | 0.0020/ 0.0015; HR 1.41 (0.62-3.21; 1) | NA | NA |
| 13 | Cabrera C I et al. November, 2023. Cleveland, USA | Cumulative incidence (%); HR at 3 months matched | Flu (contemporary cohort) | NA | NA | NA | NA | NA | NA | NA | NA | 0.01/ 0.002; HR 11.1 (8.8-13.8) | NA | NA |

|  |  | cohort (95% CI) |  |  |  |  |  |  |  |  |  |  |  |  |
| --- | --- | --- | --- | --- | --- | --- | --- | --- | --- | --- | --- | --- | --- | --- |
| 14 | Donnachie E et al. September 2022. Munchen, Germany | Cumulative incidence (%), 95% CI | ILI (historical cohort) | 18-39y: 9.10 (8.75-9.45)/ 4.99 (4.52-5.45) 40-59y: 12.33 (11.94-12.72)/ 6.10 (5.56-6.63) >60y: 10.82 (10.24-11.40) / 8.63 (7.66-9.59) | NA | 18-39y: 0.13 (0.10-0.17)/ 0.06 (0.00-0.12) 40-59y: 0.44 (0.37-0.51)/0.17 (0.10-0.24) >60y: 0.98 (0.82-1.15) / 0.83 (0.56-1.11) | NA | 18-39y: 15.29 (14.80- 15.78)/ 12.58 (11.79-13.36) 40-59y: 13.93 (13.48- 14.37)/ 10.70 (9.96-11.44) >60y: 12.26 (11.55- 12.96)/ 7.93 (7.04-8.82) | NA | 18-39y: 4.32 (4.08-4.57)/ 4.15 (3.76-4.53) 40-59y: 4.82 (4.57-5.08)/ 4.77 (4.30-5.24) >60y: 2.77 (2.47-3.08)/ 3.26 (2.72-3.80) | NA | 18-39y: 4.07 (3.86-4.28)/ 1.56 (1.31-1.80) 40-59y: 3.48 (3.28-3.68)/ 1.40 (1.15-1.65) >60y: 1.50 (1.31-1.68)/ 0.65 (0.45-0.86) | NA | NA |
| 15 | Daugherty S E et al. April, 2021. Minneapolis, USA | Cumulative incidence (%); Risk difference (95% CI; p value) | Flu (historical cohort) | NA | NA | 0.72/ 0.48; 0.24 (0.13-0.34; <0.01) | NA | 4.84/ 3.63; 1.20 (0.92-1.50; <0.01) | 0.77/ 0.78; 0.01 (-0.13-0.10; NA) | 4.53/ 4.23; 0.30 (-0.08-0.66; NA) | NA | 0.22/ 0.04; 0.18 (0.14-0.23; <0.01) | NA | NA |
| 16 | Fung KW et al. April, 2023. Bethesda, USA | Cumulative incidence (%); Adjusted OR (95% CI; p value) | ILI (historical cohort) | Outpatient: 28.0/ 24.0; OR 1.25 (1.23-1.27; <0.01) Inpatient: 43.0/31.0; OR 1.51(1.45- 1.57; <0.01) | Outpatient: 18.0/ 18.0; OR 0.97 (0.95-0.99; <0.01) Inpatient: 17.0/ 18.0; OR 0.91 (0.87-0.95; <0.01) | Memory problem Outpatient: 4.0/3.0; OR 1.09 (1.05-1.13; <0.01) Inpatient: 3.0/ 4.0; OR 0.91 (0.83-1.00; <0.01) Cognitive impairment: | Outpatient: 20.0/ 26.0; OR 0.69(0.67- 0.70; <0.01) Inpatient: 20.0/ 25.0; OR 0.69 (0.66-0.72; <0.01) | Outpatient: 33.0/ 30.0; OR 1.15(1.13-1.16; <0.01) Inpatient: 35.0/ 32.0; OR 1.25(1.21-1.30, <0.01) | Outpatient: 8.0/ 9.0; OR 0.89(0.87-0.91; <0.01) Inpatient: 8.0/ 5.0; OR 0.71 (0.66- 0.76; <0.01) | Outpatient: 6.0/ 7.0; OR 0.91(0.88-0.93; <0.01) Inpatient: 4.0/ 4.0; OR 0.86(0.79- 0.94) | Outpatient: 13.0/ 11.0; OR 1.20(1.18-1.23; <0.01) Inpatient: 14.0/ 12.0; OR 1.22(1.15- 1.28; <0.01) | Outpatient: 0.7/ 0.3; OR 2.02 (1.81-2.26; <0.01) Inpatient: 0.5/ 0.3; OR 2.04 (1.47-2.83; <0.01) | Outpatient: 1.0/ 0.9; OR 1.10 (1.02-1.18) Inpatient: 0.9/ 0.8; OR 1.09 (0.90-1.33) | NA |
|  |  |  |  |  |  | Outpatient: 3.0/<br>2.0; OR<br>1.29(1.2<br>3- 1.35;<br><0.01)<br>Inpatient<br>: 2.0/<br>2.0; OR<br>0.89(0.7<br>9- 1.01;<br><0.01) |  |  |  |  |  |  |  |  |
| 17 | Wee E L et al.<br>April, 2025.<br>Singapore,<br>Singapore | Cumulative<br>incidence<br>(%);<br>Adjusted HR<br>for outcome<br>in RSV<br>(95% CI) | COVID<br>and Flu<br>(mixed<br>cohort) | NA | NA | RSV:1.6<br>COVID:<br>1.4; HR<br>0.96<br>(0.64-1.4<br>5)<br>Flu: 0.8;<br>HR 1.34<br>(0.85-2.1<br>3) | NA | RSV:0.6<br>COVID: 0.7;<br>HR 0.76<br>(0.41-1.43)<br>Flu: 0.3; HR<br>1.32<br>(0.64-2.70) | RSV:0<br>COVID:<br>0.2; NA<br>Flu: 0.2;<br>NA | NA | NA | 0.47/<br>0.37 | NA | RSV: 0.6<br>COVID:<br>0.5; HR<br>1.33<br>(0.71-2.50<br>)<br>Flu: 0.3;<br>HR 1.60<br>(0.78-3.27<br>) |

### Occurrence of post-acute conditions after COVID-19

Post-acute conditions also varied in frequency and magnitude. Cerebrovascular disorders were consistently elevated across 14 studies, while anxiety or depression were reported in 11 studies. Seizure or epilepsy and peripheral neuropathy were each reported in 9 studies, with increased incidence after COVID-19. Cardiovascular outcomes, including coronary heart disease and heart rate abnormalities, showed modest risk increases. Type 2 diabetes mellitus was reported in eight studies, with HRs around 1.2–1.6, and pulmonary embolism and respiratory failure showed marked increases in some cohorts. Less frequently reported outcomes, such as movement and myoneural disorders, showed heterogeneous estimates. Overall, neuropsychiatric, cerebrovascular, and cardiometabolic complications were the most consistently elevated following COVID-19 (table 3).

**Table 3.** Occurrence of selected post-acute conditions in COVID-19 and comparison groups, by study.

|  | Study | Estimates | Comparison Groups Description | Post-acute conditions |  |  |  |  |  |  |  |  |  |  |
| --- | --- | --- | --- | --- | --- | --- | --- | --- | --- | --- | --- | --- | --- | --- |
|  |  |  |  | Anxiety/depression | Cerebrovascular disorders | Coronary Heart Disease | Diabetes mellitus type 2 | Heart Rate Abnormalities | Movement disorders | Myoneural Disorders | Peripheral neuropathy | Pulmonary embolism | Respiratory Failure | Seizure/epilepsy |
| 1 | Liu, Ting-Hui et al. December, 2023. Tainan, Taiwan | Cumulative incidence (%); HR (95% CI; p value) | Flu (contemporary cohort) | 7.36/6.61; HR 1.12 (0.95-1.32; 0.19) | NA | NA | NA | NA | NA | NA | NA | NA | NA | NA |
| 2 | Xie Y, Choi T, Al-Aly Z. December, 2023. Saint Louis, USA | Incident rate per 100 person at 540 days; HR (95% CI). | ILI (historical cohort) | Anxiety disorders: 15.88/ 11.82; HR 1.37 (1.29- 1.46)<br>Depression disorders: 18.12/ 15.03; HR 1.23 (1.16 -1.3) | Central Venous Thrombosis (CVT): 0.02/ 0.06; HR 0.40 (0.16 – 1.02)<br>Ischemic stroke: 8.00/ 4.74; HR 1.72 (1.56- 1.89)<br>Transient ischemic attack (TIA): 3.22/ 2.61; HR | Acute coronary disease (ACD): 14.00/ 12.42; HR 1.14 (1.07- 1.21)<br>Acute Coronary Syndrome (ACS): 4.75/ 5.48; | 11.53/ 8.48; HR 1.38 (1.29 -1.48) | Atrial fibrillation (AF): 13.61/ 11.54; HR 1.19 (1.12- 1.27)<br>Atrial flutter 1.95/ 1.90; HR 1.03 (0.88- 1.20)<br>Bradycardia: 9.75/ 6.82; HR 1.45 | NA | NA | 9.24/ 8.32; HR 1.12 (1.04- 1.2) | 6.32/ 2.82; HR 2.28 (2.02-2. 57) | NA | 4.51/ 3.41; HR 1.33 (1.19- 1.49) |
|  |  |  |  |  | 1.24<br>(1.09-1.41) | HR 0.86<br>(0.79-0.95)<br>Myocardial<br>infarction (MI):<br>12.13/<br>10.50;<br>HR 1.17<br>(1.09-1.24) |  | (1.34-1.57)<br>)<br>Tachycardia: 12.08/<br>13.62; HR<br>0.88 (0.83-0.93)<br>Ventricular<br>arrhythmias: 7.46/<br>6.20; HR<br>1.21<br>(1.11-1.32) |  |  |  |  |  |  |
| 3 | Velásquez EE et al. July, 2024. Palo Alto, USA | Cumulative incidence (%); effect size | ILI (historical cohort) | 5.2/5.7; Effect size 0.06 | 0.3/0.3; Effect size 0.02 | NA | 10.6/9.9; Effect size 0.01 | 1.6/ 1.1; Effect size 0.03 | Ataxia: 0.3/0.2; Effect size 0.01 | 0.2/0.2; Effect size 0.01 | 0.9/0.9; Effect size 0.01 | NA | NA | 0.2/ 0.2; Effect size 0.00 |
| 4 | Han, M et al. May 2024. Seoul, Korea. | Relative risk (95% CI; p value) | ILI (contemporary cohort) | 1.00 (0.12-8.33; 1.00) | NA | NA | NA | 1.27 (0.58-2.87 ; p > 0.5) | NA | NA | NA | NA | Acute: 2.50 (0.54-17.45, 0.3) | 0.15 (0.02-0.56, <0.05) |
| 5 | Whittaker H R et al. Nov 2021. London UK | Adjusted Hazard ratios (95% CI) for difference in event rates after index date compared with the same patient | COVID outpatient/ COVID inpatient/ ILI outpatient (historical cohort) | Anxiety disorders: 1.06 (1.01-1.11)/ 1.74 (1.38-2.19)/ 0.96 (0.81-1.13)<br>Depressive disorders: 1.06 (1.00-1.12)/ 1.39 (1.08-1.80) / 1.06 (0.88-1.27) | 1.08 (0.9-1.3)/ 2.49 (1.73-3.59)/ 0.96 (0.64-1.45) | MI: 0.82 (0.71-0.95)/ 2.47 (1.72-3.56)/ 1.05 (0.77-1.42) | 1.13 (1.06-1.21)/ 2.10 (1.76-2.50)/ 0.97 (0.79-1.19) | NA | NA | NA | NA | NA | NA | NA |
|  |  | 12 months before |  |  |  |  |  |  |  |  |  |  |  |  |
| 6 | Tesch F et al. October, 2024. Dresden, Germany | Cumulative incidence (%); Incidence rate ratio (95% CI) 12-15m after index date | Flu (historical cohort) | NA | NA | NA | NA | NA | NA | NA | NA | 0.22/0.09; IRR 2.32 (1.89–2.85) | Acute: 0.59/0.35 IRR: 1.51 (1.34–1.72) | NA |
| 7 | Wee L E et al. Dec, 2023. Singapore, Singapore | Cumulative incidence (%); Adjusted HR (95% CI; p value) | ILI (contemporary- Delta period and Omicron period) | Anxiety disorders: - Delta: 0.11/0.12; HR 0.84 (0.68-1.04) - Omicron: 0.11/0.13; HR 0.85(0.75-0.96; <0.05) Depression disorders: - Delta: 0.09/0.11; HR 0.80 (0.64-1.01) - Omicron: 0.05/ 0.03; HR 0.86 (0.76-0.98; <0.05) | Cerebrovascular disorders: - Delta: 0.27/ 0.18; HR 1.16 (1.00-1.33) - Omicron: 0.22/0.19; HR 0.99 (0.91-1.09) Ischemic stroke - Delta: 0.15/ 0.10; HR 1.10 (0.91- 1.32) - Omicron: 0.13/ 0.11; HR 0.98 (0.87-1.10) Hemorrhagic stroke: - Delta: 0.08/ 0.05 HR | NA | NA | NA | Delta: 0.05/0.05; 0.86 (0.63-1.17) Omicron: 0.06/0.05; HR 1.06 (0.89-1.26) | Delta: 0/0, NA Omicron: 0/0; HR 2.30 (0.97-5.44) | Delta: 0.06/0.05; HR 1.10 (0.82-1.48) Omicron: 0.05/0.05; HR 0.93 (0.77-1.11) | NA | NA | Delta: 0.03/0.02; HR 0.96 (0.61-1.51) Omicron: 0.03/ 0.03; HR 1.09 (0.86- 1.38) |
|  |  |  |  |  | 1.13(0.88-1.46)<br>- Omicron:<br>0.06/ 0.06;<br>HR 1.00<br>(0.84-1.18)<br>TIA<br>- Delta:<br>0.06/ 0.05;<br>HR 1.13<br>(0.84-1.52)<br>- Omicron:<br>0.06/ 0.05;<br>HR 1.09<br>(0.91-1.30) |  |  |  |  |  |  |  |  |  |
| 8 | Havenon A et al.<br>April,2024. New Haven, USA | Cumulative incidence (%); HR (95% CI; p value) | Flu (historical cohort) | NA | 1.97/ 2.40; HR 0.90 (0.84–0.97; <0.01) | NA | NA | NA | 1.46/ 2.46; HR 0.64 (0.60–0.69; <0.01) | NA | 1.89/ 3.61; HR 0.567 (0.53–0.60. <0.01) | NA | NA | 1.58/ 2.13; HR 0.78 (0.73–0.84; <0.01) |
| 9 | Liu TH et al.<br>November, 2023. Tainan, Taiwan | Cumulative incidence (%); HR (95% CI) | ILI (contemporary cohort) | 3.9/ 3.09; HR 1.26 (1.04-1.52) | NA | NA | NA | NA | NA | NA | NA | NA | NA | NA |
| 10 | Al-Aly Z et al.<br>April,2021. Saint Louis, USA | Incident rate per 1000 person-years; HR (95% CI, p value) | ILI (historical cohort) | 58.10/ 44.59; HR 1.31 (1.15- 1.50; 1.0) | Ischemic stroke: 18.76/ 14.58; HR 1.29 (1.03-1.61; <0.05)<br>Hemorrhagic stroke: 3.98/ 2.12; HR 1.88 (1.11-3.18; < 0.05) | MI: 23.70/ 21.29; HR 1.12 (0.92-1.35; <0.5) | 61.73/ 55.86; HR 1.11 (0.96-1.28; < 0.5) | 85.92/ 78.40; HR 1.10 (0.99-1.23; <0.1) | NA | 9.19/ 1.80; HR 5.14 (3.05-8.65; 1.0) | 35.79 / 36.72; HR 0.97 (0.83-1.14; 1.0) | 27.39 / 9.09; HR 3.04 (2.39-3.88; 1.0) | Acute: 126.41/ 56.03; HR 2.34 (2.12-2.59; 1.0) | 13.68/ 10.49; HR 1.31 (1.00-1.70; <0.05) |
|  |  |  |  |  | TIA: 5.53/<br>6.50; HR<br>0.85 (0.59-1.23; <0.5) |  |  |  |  |  |  |  |  |  |
| 11 | Baskett W et al.<br>December, 2022.<br>Columbia, USA | Cumulative<br>incidence (%);<br>OR<br>(95%<br>CI, p<br>value) | ILI<br>(contemporary cohort) | 31.7/ 31.8; OR<br>0.98<br>(0.93–1.04;<br>0.55 | Ischemic<br>stroke: 2.70/<br>2.50; OR<br>1.02 (0.87-1.20; 1.0)<br>Hemorrhagic<br>stroke:<br>0.10/ 0.20;<br>OR 1.02<br>(0.54–1.93;<br><1.0) | MI: 3.3/<br>3.1; OR<br>1.02<br>(0.87-1.2;<br><1.0)<br>ACS:<br>0.20/<br>0.20;<br>OR 0.89<br>(0.51-1.<br>55;<br><1.0) | 21.9/<br>20.6;<br>OR<br>1.19<br>(1.08–<br>1.32;<br><0.00<br>5) | AF: 7.1/<br>6.9; OR<br>1.04<br>(0.9-1.2;<br><1.0)<br>Tachycardia:<br>5.2/<br>4.3; OR<br>1.28<br>(1.15-1.42<br>; <0.01) | NA | NA | 3.8/ 3.5;<br>OR 1.14<br>(1.0–1.3<br>1;<br><0.05) | 1.1/ 0.9;<br>OR<br>1.36<br>(1.07–1.<br>74;<br><0.05) | 5.2/ 4.5;<br>OR 1.27<br>(1.12–1.<br>44;<br><0.01) | NA |
| 12 | Cohen K et al.<br>December, 2021.<br>Minnetonka, USA | Incident<br>rates per<br>person-years; HR<br>(95%<br>CI, p<br>value) | ILI<br>(historical cohort) | NA | Ischemic<br>stroke:<br>0.067/<br>0.074; HR<br>0.88 (0.77-1.01; 1.0)<br>Hemorrhagic<br>stroke:<br>0.045/<br>0.051; HR<br>0.85 (0.72-1.00; 1.0) | MI:<br>0.0414/<br>0.055;<br>HR 0.72<br>(0.62-0.85;<br>1.0)<br>ACS:<br>0.049/<br>0.067;<br>HR 0.71<br>(0.61-0.83;<br>1.0) | 0.094<br>9/<br>0.075<br>8; HR<br>1.23<br>(1.02-1.48,<br><0.00<br>5) | 0.1475/<br>0.1585;<br>HR 0.92<br>(0.83-1.01<br>; 1.0) | NA | NA | 0.0284/<br>0.0354;<br>HR 0.79<br>(0.65-0.96;<br>1.0) | 0.0418/<br>0.0335;<br>HR<br>1.19<br>(0.99-1.42;<br><0.05) | Acute:<br>0.2887/<br>0.1503;<br>HR 1.9<br>(1.75-2.05;<br><0.01)<br>Chronic:<br>0.0431/<br>0.0512;<br>HR 0.82<br>(0.69-0.<br>96; 1.0) | 0.0176/<br>0.018); HR<br>0.95<br>(0.74-1.24;<br>1.0) |
| 13 | Donnachie E et al.<br>September 2022.<br>Munich, Germany | Cumulative<br>incidence (%),<br>95% CI | ILI<br>(historical cohort) | NA | NA | NA | NA | NA | NA | NA | NA | 18-39y:<br>0.17<br>(0.12-0.22)/<br>0.12<br>(0.04-0.20)<br>40-59y:<br>0.50<br>(0.43-0.56)/<br>0.29 | NA | NA |
|  |  |  |  |  |  |  |  |  |  |  |  | (0.19-0.38)<br>>60y:<br>0.95<br>(0.80-1.09)/<br>0.75<br>(0.49-1.01) |  |  |
| 14 | Daugherty S E et al.<br>April, 2021.<br>Minneapolis, USA | Cumulative incidence (%); Risk difference (95% CI; p value) | Flu (historical cohort) | Anxiety: 3.08/2.48; 0.60 (0.32- 0.84; <0.01)<br>Depression: 1.65/ 1.63; 0.03 (-0.18-0.20) | Stroke: 0.22/ 0.18; 0.05 (-0.02-0.10)<br>Ischemic stroke: 0.19/ 0.15; 0.04 (-0.01- 0.09)<br>Hemorrhagic stroke: 0.06/ 0.05; 0.01 (-0.02 -0.04) | ACS: 0.20/ 0.21; -0.01 (-0.08-0.05)<br>MI: 0.14/ 0.13; 0.01 (-0.04-0.06)<br>ACD: 0.22/ 0.23; -0.01 (-0.07-0.06) | 1.05/ 0.77; 0.28 (0.13-0.41; <0.01) | Arrhythmia : 1.52/ 1.09; 0.44 (0.26-0.62 ; <0.01)<br>Cardiac arrhythmia: 0.72/ 0.51; 0.21 (0.10-0.31 )<br>Tachycardia: 1.11/ 0.77; 0.34 (0.20-0.48; <0.01) | NA | NA | 0.32/ 0.20; 0.12 (0.05-0.20; <0.01) | 0.20/ 0.13; 0.07 (0.02-0.12; <0.01) | Respiratory Failure: 0.26/ 0.27; -0.04 (-0.11-0.02)<br>Acute: 0.22/ 0.26; -0.04 (-0.10-0.03)<br>Chronic: 0.18/ 0.11; 0.07 (0.02-0.12; <0.01) | 0.14/ 0.14; -0.00 (-0.05-0.04) |
| 15 | Wee E L et al.<br>April, 2025.<br>Singapore, Singapore | Cumulative incidence (%); Adjusted HR (95% CI) for outcome in RSV compared with | FLU and RSV (mixed cohort) | NA | RSV:0.8<br>COVID: 0.9; HR 0.79 (0.46-1.38)<br>Flu: 0.7; HR 0.81 (0.45-1.47) | RSV:2.0<br>COVID: 1.4; HR 1.24 (0.86-1.77)<br>Flu: 1.3; HR 1.18 (0.79-1.76) | RSV:2.3<br>COVID: 2.0; HR 0.93 (0.66-1.30)<br>Flu: 2.0; HR 0.85 | RSV:1.5<br>COVID: 1.3; HR 1.03 (0.68-1.56)<br>Flu: 1.2; HR 0.97 (0.62-1.52) | RSV:0.3<br>COVID : 0.3; HR 0.93 (0.35-2.52)<br>Flu: 0.1; HR 2.24 (0.69-7.28) | NA | RSV:0.5<br>COVID : 0.2; HR 2.19 (1.07-4.49)<br>Flu: 0.2; HR 1.74 (0.77-3.97) | NA | NA | NA |
|  |  | COVID and Flu |  |  |  |  | (0.59-1.22) |  |  |  |  |  |  |  |
| 16 | Quinn K L et al. June, 2023. Toronto, Canada | Crude rate per 100 person-year; HR (95% CI) | ILI (historical cohort) | NA | 1.30 / 2.48; HR 0.70 (0.57- 0.86) | MI: 0.07/ 0.14; HR 1.00 (0.47- 2.12) | NA | NA | NA | NA | NA | NA | NA | 0.14/ 0.28; HR 0.54 (0.30- 0.95) |
| 17 | Xie Y et al. January, 2022. Saint Louis, USA | Hazard ratio (95% CI) | Flu (historical cohort) | Anxiety inpatient: 1.34 (1.13-1.59)<br>Anxiety outpatient: 1.44 (1.22-1.71)<br>Depression inpatient: 1.24 (1.06-1.47)<br>Depression outpatient: 1.32 (1.15-1.56) | NA | NA | NA | NA | NA | NA | NA | NA | NA | NA |
| 18 | Zarifkar P et al. June, 2022. Copenhagen, Denmark | Cumulative incidence (%); RR (95% CI) | Flu (historical cohort) | NA | Ischemic stroke: 0.64/ 0.80; RR 0.80 (0.60- 1.10)<br>Hemorrhagic stroke: 0.04/ 0.01; N/A | NA | NA | NA | NA | NA | NA | NA | NA | NA |
| 19 | Ward A et al. January, 2022. California, USA | Cumulative incidence (%); HR (95% CI) | ILI (historical cohort) | NA | 0.84/ 0.35; 1.11 (0.98- 1.25) | MI: 0.98/ 0.58; 0.93 (0.85- 1.03) | NA | NA | NA | NA | NA | 0.55/ 0.22; 1.82 (1.57- 2.10) | NA | NA |
| 20 | Lee, H et al. Feb 2022. Seoul, South Korea | Adjusted OR (95% CI) of each age | ILI (historical cohort) | NA | 45-64 years: 5.23 (1.55-17.67) | NA | NA | NA | NA | NA | NA | NA | NA | NA |
|  |  | range<br>compare<br>d with<br>referenc<br>e<br>(20-44y) |  |  | 65-74 years:<br>7.30 (2.06-25.84)<br>>75 years:<br>9.02 ( 2.48-32.75) |  |  |  |  |  |  |  |  |  |

### Meta-analysis

Fourteen studies with compatible data were included in the meta-analysis. Across 24 evaluated outcomes, six showed significantly higher pooled risks following SARS-CoV-2 infection compared with other respiratory viral infections (influenza, ILI, or RSV) (figure 2).

**Figure 2.**
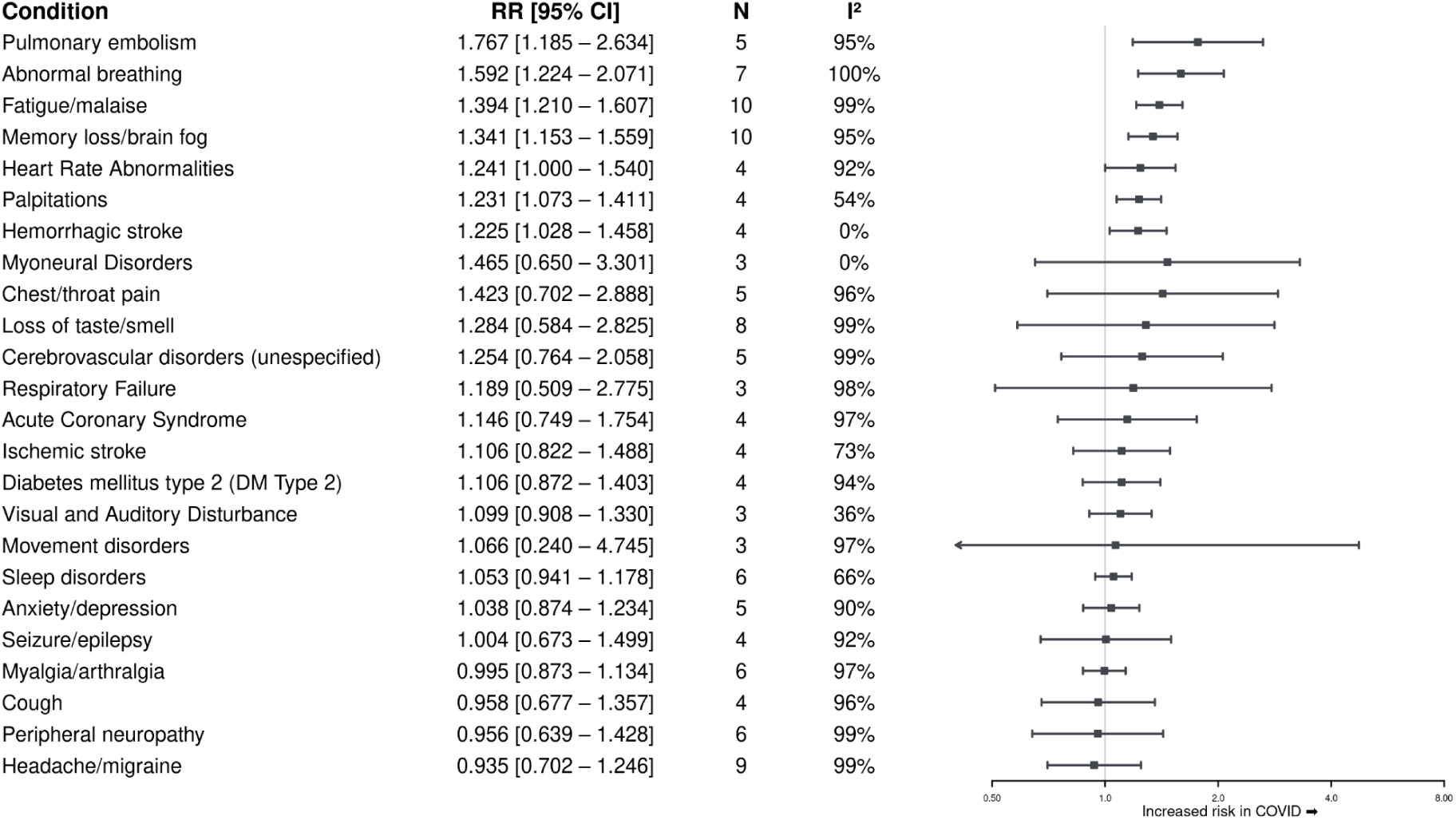
Pooled relative risks of post-acute outcomes after SARS-CoV-2 infection compared with other respiratory viral infections. **Alt text:** Forest plot summarizing pooled RRs with 95% CIs. Six conditions showed significantly higher risk after SARS-CoV-2C (pulmonary embolism, abnormal breathing, fatigue/malaise, memory loss/brain fog, palpitations, and hemorrhagic stroke), while all other outcomes were not significantly different after other viral infections.

Significantly increased risks were observed for pulmonary embolism (RR 1.767; 95% CI 1.185–2.634), abnormal breathing (RR 1.592; 95% CI 1.224–2.071), fatigue or malaise (RR 1.394; 95% CI 1.210–1.607), memory loss or brain fog (RR 1.341; 95% CI 1.153–1.559), palpitations (RR 1.231; 95% CI 1.073–1.411), and hemorrhagic stroke (RR 1.225; 95% CI 1.028–1.458). Heart rate abnormalities showed borderline significance (RR 1.241; 95% CI 1.000–1.540) (appendix pp 28–30).

No increased risk was observed for anxiety or depression, acute coronary syndrome, cerebrovascular disorders, headache or migraine, loss of taste or smell, or sleep disorders, suggesting substantial overlap between post-acute manifestations of COVID-19 and those observed after other respiratory viral infections (PAIS).

### Assessment of bias and robustness

Publication bias was assessed only for fatigue/malaise and memory loss/brain fog, the only outcomes reported in ≥10 studies. Funnel plots showed no clear asymmetry (appendix pp 31). Egger’s and Begg’s tests were not significant for fatigue/malaise (p=0.182; p=0.788) or memory loss/brain fog (p=0.0737; p=0.531), indicating no clear small-study effects.

Meta-regression analyses including study precision as a covariate showed no significant association between effect size and study precision (β = 0.14, 95% CI −0.01 to 0.29; p = 0.067), suggesting that variability across outcomes likely reflects true differences rather than publication bias (appendix p 32).

Influence diagnostics (Cook’s distance, DFFITS, covariance ratios, and standardized residuals) showed that no individual study disproportionately influenced the pooled estimates (appendix p 33).

## Discussion

In this systematic review and meta-analysis, acute SARS-CoV-2 infection was associated with an increased risk of a limited set of post-acute outcomes compared with other acute respiratory viral infections, including pulmonary embolism, abnormal breathing, fatigue, memory loss or brain fog, palpitations, heart rate abnormalities, and hemorrhagic stroke. No higher risk was observed for the remaining analysed symptoms and conditions. These findings suggest that post-acute infection syndromes may occur after non-COVID respiratory infections and share many clinical features with post-COVID condition (PCC), whereas SARS-CoV-2 infection may increase the risk of specific thrombotic, autonomic, and neurocognitive outcomes.

The increased risk of pulmonary embolism is consistent with evidence that SARS-CoV-2 promotes a multifactorial prothrombotic state, often referred to as COVID-19-associated coagulopathy or immunothrombosis.^49^ Endothelial activation and injury disrupt antithrombotic homeostasis and promote platelet aggregation,^50^ while activated immune cells release tissue factor, inflammatory cytokines, and neutrophil extracellular traps that amplify thrombogenesis.^51^ Complement activation, impaired fibrinolysis, and microvascular dysfunction may further contribute to thrombus formation.^49,52^ In addition, dysregulation of the renin-angiotensin system following viral binding to the angiotensin-converting enzyme 2 receptor may promote vasoconstriction, endothelial dysfunction, and vascular fragility, potentially increasing the risk of hemorrhagic cerebrovascular events after infection.^54,55^

Several non-thrombotic outcomes observed in our analysis may be explained by mechanisms similar to those described in myalgic encephalomyelitis or chronic fatigue syndrome. Immune dysregulation, persistent neuroinflammation, and viral reactivation may contribute to prolonged fatigue and cognitive dysfunction.^51^ Mitochondrial impairment, redox imbalance, and endothelial or microvascular dysfunction may reduce oxygen delivery and utilisation, providing a biological basis for exercise intolerance and abnormal breathing.^56–58^ Autonomic nervous system involvement, including postural orthostatic tachycardia syndrome, has been reported both in ME/CFS and PCC and may underlie palpitations and heart rate abnormalities.^59–60^

Previous systematic reviews of uncontrolled studies have reported a high prevalence of persistent symptoms after COVID-19.^62^ However, many symptoms commonly attributed to PCC were not significantly increased in our analyses when compared with viral or influenza-like illness control populations. These findings highlight the importance of appropriate comparator groups when attempting to identify symptoms that are truly specific to SARS-CoV-2 infection, particularly for non-specific symptoms that are common in the general population or influenced by broader pandemic-related factors.

Meta-analyses comparing SARS-CoV-2 infection with uninfected controls have generally reported higher relative risks for a wide range of symptoms, including fatigue, breathlessness, and cognitive impairment.^63^ Although the direction of associations for fatigue and cognitive outcomes was consistent with our findings, effect sizes were generally larger when uninfected controls were used. Similarly, large meta-analyses including more than 14 million participants have reported increased risks for numerous long-COVID symptoms, with particularly high estimates for anosmia, ageusia, and cognitive outcomes.^14^ The lower effect estimates observed in our analyses may reflect the use of symptomatic comparator groups, which likely capture post-viral sequelae unrelated specifically to SARS-CoV-2.

Evidence from paediatric populations also supports an overlap between PCC and other post-viral syndromes. Comparative studies in children have reported similar symptom profiles and duration between post-COVID and other post-viral conditions, reinforcing the concept of shared post-infectious mechanisms across different pathogens.^64^

Differences in risk estimates across studies and meta-analyses are likely multifactorial. High heterogeneity between studies, differences in patient populations (hospitalized and outpatients), variation in follow-up duration, and the lack of data on viral variants or vaccination status complicate interpretation. Although the studies included in our meta-analysis were generally of high methodological quality according to the Newcastle-Ottawa Scale, substantial between-study heterogeneity persisted for several outcomes.

This study has several limitations. Most included studies were retrospective, limiting causal inference and introducing potential misclassification and residual confounding, particularly for subjective symptoms assessed long after the acute infection. The absence of stratified data by vaccination status, viral variant, timing of post-acute outcome, and clinical care setting also limits the generalisability of the findings to later phases of the pandemic.

A key strength of this meta-analysis is the use of symptomatic comparator populations. Comparing post-COVID outcomes with individuals experiencing other viral respiratory infections provides a more appropriate reference group than uninfected controls and helps distinguish SARS-CoV-2–specific sequelae from broader post-viral syndromes.

Overall, our findings suggest that several symptoms frequently attributed to PCC may reflect shared post-infectious pathways rather than SARS-CoV-2–specific effects. Distinguishing pathogen-specific sequelae from common post-viral syndromes has important implications for patient counselling, risk stratification, health-care planning, and the design of post-infectious surveillance and rehabilitation strategies. Future research should prioritise large prospective comparator-based cohort studies with harmonised outcome definitions and stratification by vaccination status and viral variant to clarify pathogen-specific mechanisms and inform evidence-based care.

## Acknowledgements

Ethics statement

This study is based on previously published data and does not require ethical approval.

## Data availability

All data used in this study are available in the original published articles.

## Author’s contributions

TFP: conceptualisation, investigation, data curation, software, methodology, formal analysis, visualisation, writing – original draft and writing– review & editing.

AS: investigation, data curation, software, validation

ALGO: methodology, data curation, software, formal analysis, supervision, writing – original draft

AA: investigation, data curation, validation, writing– review & editing

TST:methodology, formal analysis, software, visualization, writing – original draft

FI: funding acquisition, project administration, resources, supervision, writing– review & editing

GM: investigation, supervision, writing– review & editing

ACL: methodology, supervision, writing– review & editing

JP: conceptualisation, funding acquisition, project administration, resources, writing–review & editing

JFMC: conceptualisation, investigation, data curation, formal analysis, methodology, project administration, supervision, validation, visualisation, writing – original draft, and writing– review & editing.

## Declaration of generative AI and AI-assisted technologies in the manuscript preparation process

During the preparation of this work the authors used ChatGPT (OpenAI, San Francisco, CA, USA) to assist with language editing and text condensation. After using this tool/service, the authors reviewed and edited the content as needed and take full responsibility for the content of the published article.

