## Supplementary appendix for "Risk of Post-acute Symptoms and Conditions After SARS-CoV-2 Compared to Other Respiratory Viral Infections: A Systematic Review and Meta-Analysis"

### List of Contents


Table S1. PRISMA Checklist.

Table S2. PECO strategy.

Table S3. Outcomes included in the Core Outcome Set - PC-COS.

Table S4. Quality assessment of included studies (Newcastle–Ottawa Scale).

Table S5. Outcome Coding Based on the PCC Core Outcome Set (International Delphi Consensus).

Table S6. Outcome harmonization - symptoms.

Table S7. Outcome harmonization - conditions.

Table S8. Post-acute outcomes included in the meta-analysis, by study.

Figure S1. Pooled risk of pulmonary embolism (COVID vs ILI).

Figure S2. Pooled risk of abnormal breathing (COVID vs ILI).

Figure S3. Pooled risk of fatigue/malaise (COVID vs ILI).

Figure S4. Pooled risk of memory loss/brain fog (COVID vs ILI).

Figure S5. Pooled risk of heart Rate Abnormalities (COVID vs ILI).

Figure S6. Pooled risk of palpitations (COVID vs ILI).

Figure S7. Pooled risk of hemorrhagic stroke (COVID vs ILI).

Figure S8. Funnel plots for outcomes with ≥10 studies.

Figure S9. Overall funnel plot including all conditions.

Figure S10. Meta-regression coefficient plot adjusted for study precision.

Figure S11. Relationship between study precision and effect size.

Figure S12. Influence diagnostics and sensitivity analyses.

**Table S1.** PRISMA Checklist.

| Section and Topic | Item # | Checklist item | Location where item is reported |
|---|---|---|---|
| **TITLE** | | | |
| Title | 1 | Identify the report as a systematic review. | Title |
| **ABSTRACT** | | | |
| Abstract | 2 | See the PRISMA 2020 for Abstracts checklist. | Abstract |
| **INTRODUCTION** | | | |
| Rationale | 3 | Describe the rationale for the review in the context of existing knowledge. | Introduction |
| Objectives | 4 | Provide an explicit statement of the objective(s) or question(s) the review addresses. | Introduction |
| **METHODS** | | | |
| Eligibility criteria | 5 | Specify the inclusion and exclusion criteria for the review and how studies were grouped for the syntheses. | Methods, appendix |
| Information sources | 6 | Specify all databases, registers, websites, organisations, reference lists and other sources searched or consulted to identify studies. Specify the date when each source was last searched or consulted. | Methods |
| Search strategy | 7 | Present the full search strategies for all databases, registers and websites, including any filters and limits used. | Methods, appendix |
| Selection process | 8 | Specify the methods used to decide whether a study met the inclusion criteria of the review, including how many reviewers screened each record and each report retrieved, whether they worked independently, and if applicable, details of automation tools used in the process. | Methods |
| Data collection process | 9 | Specify the methods used to collect data from reports, including how many reviewers collected data from each report, whether they worked independently, any processes for obtaining or confirming data from study investigators, and if applicable, details of automation tools used in the process. | Methods, appendix |
| Data items | 10a | List and define all outcomes for which data were sought. Specify whether all results that were compatible with each outcome domain in each study were sought (e.g. for all measures, time points, analyses), and if not, the methods used to decide which results to collect. | Methods, appendix |

| | | | |
|---|---|---|---|
| | 10b | List and define all other variables for which data were sought (e.g. participant and intervention characteristics, funding sources). Describe any assumptions made about any missing or unclear information. | Methods, appendix |
| Study risk of bias assessment | 11 | Specify the methods used to assess risk of bias in the included studies, including details of the tool(s) used, how many reviewers assessed each study and whether they worked independently, and if applicable, details of automation tools used in the process. | Methods, appendix |
| Effect measures | 12 | Specify for each outcome the effect measure(s) (e.g. risk ratio, mean difference) used in the synthesis or presentation of results. | Methods, results, appendix |
| Synthesis methods | 13a | Describe the processes used to decide which studies were eligible for each synthesis (e.g. tabulating the study intervention characteristics and comparing against the planned groups for each synthesis (item #5)). | Methods, appendix |
| | 13b | Describe any methods required to prepare the data for presentation or synthesis, such as handling of missing summary statistics, or data conversions. | Methods, appendix |
| | 13c | Describe any methods used to tabulate or visually display results of individual studies and syntheses. | Methods, appendix |
| | 13d | Describe any methods used to synthesize results and provide a rationale for the choice(s). If meta-analysis was performed, describe the model(s), method(s) to identify the presence and extent of statistical heterogeneity, and software package(s) used. | Methods, appendix |
| | 13e | Describe any methods used to explore possible causes of heterogeneity among study results (e.g. subgroup analysis, meta-regression). | Methods, appendix |
| | 13f | Describe any sensitivity analyses conducted to assess robustness of the synthesized results. | Methods, appendix |
| Reporting bias assessment | 14 | Describe any methods used to assess risk of bias due to missing results in a synthesis (arising from reporting biases). | Methods, appendix |
| Certainty assessment | 15 | Describe any methods used to assess certainty (or confidence) in the body of evidence for an outcome. | Methods, appendix |
| **RESULTS** | | | |
| Study selection | 16a | Describe the results of the search and selection process, from the number of records identified in the search to the number of studies included in the review, ideally using a flow diagram. | Results |
| | 16b | Cite studies that might appear to meet the inclusion criteria, but which were excluded, and explain why they were excluded. | NA |

| | | | |
|---|---|---|---|
| Study characteristics | 17 | Cite each included study and present its characteristics. | Methods, results |
| Risk of bias in studies | 18 | Present assessments of risk of bias for each included study. | Methods, results, appendix |
| Results of individual studies | 19 | For all outcomes, present, for each study: (a) summary statistics for each group (where appropriate) and (b) an effect estimate and its precision (e.g. confidence/credible interval), ideally using structured tables or plots. | Methods, results, appendix |
| Results of syntheses | 20a | For each synthesis, briefly summarise the characteristics and risk of bias among contributing studies. | Methods, results, appendix |
| | 20b | Present results of all statistical syntheses conducted. If meta-analysis was done, present for each the summary estimate and its precision (e.g. confidence/credible interval) and measures of statistical heterogeneity. If comparing groups, describe the direction of the effect. | Results, appendix |
| | 20c | Present results of all investigations of possible causes of heterogeneity among study results. | Results, appendix |
| | 20d | Present results of all sensitivity analyses conducted to assess the robustness of the synthesized results. | Results, appendix |
| Reporting biases | 21 | Present assessments of risk of bias due to missing results (arising from reporting biases) for each synthesis assessed. | Results, appendix |
| Certainty of evidence | 22 | Present assessments of certainty (or confidence) in the body of evidence for each outcome assessed. | Results, appendix |
| **DISCUSSION** | | | |
| Discussion | 23a | Provide a general interpretation of the results in the context of other evidence. | Discussion |
| | 23b | Discuss any limitations of the evidence included in the review. | Discussion |
| | 23c | Discuss any limitations of the review processes used. | Discussion |
| | 23d | Discuss implications of the results for practice, policy, and future research. | Discussion |
| **OTHER INFORMATION** | | | |

| | | | |
|---|---|---|---|
| Registration and protocol | 24a | Provide registration information for the review, including register name and registration number, or state that the review was not registered. | Methods |
| | 24b | Indicate where the review protocol can be accessed, or state that a protocol was not prepared. | Methods |
| | 24c | Describe and explain any amendments to information provided at registration or in the protocol. | NA |
| Support | 25 | Describe sources of financial or non-financial support for the review, and the role of the funders or sponsors in the review. | Abstract, methods |
| Competing interests | 26 | Declare any competing interests of review authors. | Declaration of interests |
| Availability of data, code and other materials | 27 | Report which of the following are publicly available and where they can be found: template data collection forms; data extracted from included studies; data used for all analyses; analytic code; any other materials used in the review. | Methods, results, appendix |

NA: Not applied

*From:* Page MJ, McKenzie JE, Bossuyt PM, Boutron I, Hoffmann TC, Mulrow CD, et al. The PRISMA 2020 statement: an updated guideline for reporting systematic reviews. BMJ 2021;372:n71. doi: 10.1136/bmj.n71 For more information, visit: http://www.prisma-statement.org/

### Table S2. PECO strategy.

| Elements | Description / Terms used |
|---|---|
| **P (Population)** | Young Adult OR Middle Aged OR Aged |
| **E (Exposition)** | COVID-19 or SARS-CoV-2 |
| **C (Comparator group)** | Influenza A Virus, H1N1 Subtype OR Influenza, Human OR Influenza B virus OR Respiratory Syncytial Viruses OR Pneumovirus OR Human adenovirus 1 OR Adenoviruses, Human OR Rhinovirus OR Human rhinovirus OR JC Virus OR Metapneumovirus OR Human metapneumovirus OR Parotitis OR Rubella virus OR Morbillivirus |
| **O (Outcomes)** | complications OR sequelae OR convalesc* OR Fatigue Syndrome, Chronic OR Dyspnea OR cognitive dysfunction OR long-covid OR PASC |
| **T (Type of study)** | Observational studies, Longitudinal, Cohort, Prospective cohort, Retrospective matched cohort, Retrospective cohort, Cross-sectional |

### Search Strategy Elements

07 Jun 2024

| Database | Search Description | Results |
|---|---|---|
| **PubMed** | (((((((((((((((((("Young Adult"[Mesh]) OR "Middle Aged"[Mesh]) OR "Aged"[Mesh]) AND "SARS-CoV-2"[Mesh]) AND "Influenza, Human"[Mesh]) OR "Influenza A Virus, H1N1 Subtype"[Mesh]) OR "Influenza B virus"[Mesh]) OR "Respiratory Syncytial Virus, Human"[Mesh]) OR "Adenoviruses, Human"[Mesh]) OR "Rhinovirus"[Mesh]) OR "Metapneumovirus"[Mesh]) OR "Parainfluenza Virus 1, Human"[Mesh]) OR "Parainfluenza Virus 2, Human"[Mesh]) OR "Parainfluenza Virus 3, Human"[Mesh]) OR "Parainfluenza Virus 4, Human"[Mesh]) AND "complications" [Subheading]) OR "Convalescence"[Mesh]) OR "sequelae" OR "Post-Acute COVID-19 Syndrome"[Mesh]) | **6838** |
| **Scopus** | ( ALL ( "Young Adult" OR "Middle Aged" OR aged ) AND ALL ( "sars cov 2" ) AND ALL ( "Influenza, Human" OR "Influenza B virus" OR "influenza B" OR "Respiratory Syncytial Viruses" OR pneumovirus OR "Human adenovirus 1" OR "Adenoviruses, Human" OR rhinovirus OR "Human rhinovirus" OR metapneumovirus OR "Human metapneumovirus" OR | **1632** |

| | | |
|---|---|---|
| | "parainfluenza virus" ) AND ALL ( "post acute sequelae of covid 19" ) OR ALL ( complications OR sequelae OR convalesc* ) ) AND ( LIMIT-TO ( DOCTYPE , "ar" ) ) | |
| **Embase** | ('young adult'/exp OR 'young adult' OR 'middle aged'/exp OR 'middle aged' OR 'aged'/exp OR 'aged') AND ('severe acute respiratory syndrome coronavirus 2'/exp OR 'severe acute respiratory syndrome coronavirus 2') AND ('influenza a virus, h1n1 subtype'/exp OR 'influenza a virus, h1n1 subtype' OR 'influenza, human'/exp OR 'influenza, human' OR 'influenza b virus'/exp OR 'influenza b virus' OR 'influenza b'/exp OR 'influenza b' OR 'respiratory syncytial viruses'/exp OR 'respiratory syncytial viruses' OR 'pneumovirus'/exp OR 'pneumovirus' OR 'human adenovirus 1'/exp OR 'human adenovirus 1' OR 'adenoviruses, human'/exp OR 'adenoviruses, human' OR 'rhinovirus'/exp OR 'rhinovirus' OR 'human rhinovirus'/exp OR 'human rhinovirus' OR 'metapneumovirus'/exp OR 'metapneumovirus' OR 'human metapneumovirus'/exp OR 'human metapneumovirus' OR 'human parainfluenza virus 1'/exp OR 'human parainfluenza virus 1' OR 'human parainfluenza virus 2'/exp OR 'human parainfluenza virus 2' OR 'human parainfluenza virus 3'/exp OR 'human parainfluenza virus 3' OR 'human parainfluenza virus 4'/exp OR 'human parainfluenza virus 4') AND ('long covid'/exp OR 'long covid' OR 'complications'/exp OR 'complications' OR sequelae OR convalesc*) | **901** |
| **Total** | | **9371** |

**Table S3. Outcomes included in the Core Outcome Set - PC-COS.**

| Domain | Outcome | Outcome description |
|---|---|---|
| Mortality | Survival | How long does someone live |
| Physiologica l/Clinical Outcomes | Cardiovascular functioning; symptoms; and conditions | New onset or worsening of problems affecting the heart (e.g. pounding or racing heart) and the blood vessels (e.g., veins or arteries) |
| | Fatigue or Exhaustion | New onset or worsening in severity or duration of feeling exhausted, having too little energy, or needing more rest |
| | Pain | New onset or worsening of problems related to uncomfortable feelings in the body that can include sharp or burning pain, dull ache, or stinging or throbbing feeling, pain that comes and goes |
| | Nervous system functioning; symptoms; and conditions | New onset or worsening of dizziness, fainting, headache, tremors/shaking, seizures/fits, muscle twitching, tingling feelings, decreased sensation, stroke, inability to move part of the body, lack of coordination, or speech difficulty |
| | Cognitive functioning; symptoms; and conditions | New onset or worsening problems with memory, communication, concentration, or understanding instructions |
| | Mental functioning; symptoms; and conditions | New onset or worsening problems with emotions and mood, including anxiety/worrying, panic attacks, depression, suicidal thoughts, or post-traumatic stress disorder |

| | | |
|---|---|---|
| | Respiratory functioning; symptoms; and conditions | New onset or worsening problems with lungs or breathing (e.g., shortness of breath, chest tightness. or coughing) |
| | Post-exertion symptoms | Worsening of symptoms following physical or mental exertion that can last for a prolonged duration |
| Life Impact Outcomes | Physical functioning; symptoms; and conditions | New onset or worsening problems with physical abilities, including muscle strength, arm/leg shaking or unsteadiness, walking, dressing, or eating |
| | Work/occupational changes and study | New onset or worsening problems with being able to resume work, study or activities/hobbies |
| **Outcomes included from the COVID-19 COS [1]** | | |
| | Recovery | The absence of symptoms related to the illness, the ability to do usual daily activities, and a return |

From: Consensus Meeting Report V0.2 2-12-2021 [17]

**Table S4. Quality assessment of included studies (Newcastle–Ottawa Scale).**

| ID | Authors | Selection | Comparability | Exposure/ Outcome | Overall |
|---|---|---|---|---|---|
| **1** | Liu et al., 2023 | 4 | 2 | 3 | 9 |
| **2** | Velásquez et al., 2024 | 4 | 2 | 3 | 9 |
| **3** | Tesch et al., 2024 | 4 | 2 | 3 | 9 |
| **4** | Wee et al., 2023 | 4 | 2 | 3 | 9 |
| **5** | Havenon et al., 2024 | 3 | 2 | 3 | 8 |
| **6** | Zarifkar et al., 2022 | 4 | 1 | 3 | 8 |
| **7** | Liu et al., 2023 | 3 | 2 | 3 | 8 |
| **8** | Baskett et al., 2022 | 3 | 2 | 3 | 8 |
| **9** | Cabrera et al., 2023 | 4 | 1 | 3 | 8 |
| **10** | Fung et al., 2023 | 4 | 2 | 2 | 8 |
| **11** | Ward et al., 2022 | 4 | 2 | 2 | 8 |
| **12** | Wee et al., 2025 | 4 | 2 | 3 | 9 |
| **13** | Donnachie et al., 2022 | 4 | 2 | 3 | 9 |
| **14** | Han et al., 2024 | 4 | 2 | 2 | 8 |
| **15** | Xie et al., 2023 | 2 | 2 | 3 | 7 |
| **16** | Quinn et al., 2023 | 4 | 2 | 3 | 9 |
| **17** | Al-Aly et al., 2021 | 3 | 2 | 3 | 8 |
| **18** | Cohen et al., 2021 | 4 | 2 | 2 | 8 |
| **19** | Daugherty et al., 2021 | 4 | 2 | 2 | 8 |
| **20** | Xie et al., 2022 | 3 | 2 | 2 | 7 |
| **21** | Lee et al., 2022 | 4 | 2 | 2 | 8 |
| **22** | Whittaker et al., 2021 | 4 | 2 | 2 | 8 |

The NOS evaluates observational studies across three parameters: selection, comparability between exposed and unexposed groups, and exposure or outcome assessment. Each domain can receive a maximum of 4, 2, and 3 stars, respectively. Studies scoring fewer than 5 stars are classified as low quality, those scoring 5–7 stars are considered moderate quality, and studies scoring more than 7 stars are categorized as high quality.

**Table S5 - Outcome Coding Based on the PCC Core Outcome Set (International Delphi Consensus).**

| |
|---|
| Physiological or clinical outcomes<br>(1) Cardiovascular functioning, symptoms and conditions<br>(2) Fatigue/exhaustion<br>(3) Pain<br>(4) Nervous system functioning, symptoms and conditions<br>(5) Cognitive functioning, symptoms and conditions<br>(6) Mental functioning, symptoms and conditions<br>(7) Respiratory functioning, symptoms and conditions<br>(8) Post exertion symptoms |
| Life impact<br>(9) Physical functioning, symptoms and conditions<br>(10) Work or occupation and study changes |
| Survival<br>(11) Survival |
| Outcome from previous COS<br>(12) Recovery |
| Other<br>(13) Other |

**Note: The numbers in brackets correspond to the codes used in Tables S6, S7 and S8.**

**Table S6. Harmonization of symptom outcomes.**

| System | Before harmonization | After harmonization |
|---|---|---|
| Cardiovascular functioning, symptoms and conditions (1) | Palpitation, palpitations | Palpitations |
| Fatigue/exhaustion (2) | Chronic Fatigue Syndrome; Fatigue; Fatigue Diagnosis (composite); Fatigue/malayse; Malaise/exhaustion; Somnolence, malaise, and fatigue; Fatigue/ malaise/ weakness | Fatigue/malaise |
| Pain (3) | Joint pain, Myalgia, Muscle/joint pain | Myalgia/arthralgia |
| Nervous system functioning, symptoms and conditions (4) | Headache, Headache disorders, Migraine | Headache/Migraine |
| | Loss of taste/smell, Loss of smell or taste, Smell and taste disorders, Anosmia | Loss of taste/smell |
| Nervous system functioning, symptoms and conditions (4) | Hearing abnormalities or tinnitus, Tinnitus, Visual abnormalities, Visual and Auditory Disturbance | Visual and Auditory Disturbance |
| Cognitive functioning, symptoms and conditions (5) | Cognitive Impairment, Cognitive Disturbance, Cognitive Dysfunction, Cognitive symptoms; Cognition and Memory disorders; Amnesia/Memory Difficulty, Memory disorder, Memory loss, Memory problems, Memory problem | Memory loss/brain fog |
| Mental functioning, symptoms and conditions (6) | Insomnia, Sleep Disturbances, Sleep disturbance, Sleep disorders | Sleep disorders |

| | | |
|---|---|---|
| Respiratory; Respiratory functioning, symptoms and conditions (7) | Abnormal breathing, Breathing Difficulties, Dyspnea, Shortness of Breath | Abnormal breathing |
| | Chest pain, Chest/throat pain | Chest/throat pain |
| | Cough | Cough |

**Note: The numbers in brackets correspond to the codes explained in Tables S5.**

**Table S7 - Harmonization of condition outcomes.**

| System | Before harmonization | After harmonization |
|---|---|---|
| Cardiovascular/ Cardiovascular function, symptoms and conditions (1) | Atrial fibrillation; Heart Rate Abnormalities; Cardiac Rhythm Disorders (composite); Cardiac Arrhythmia; Arrhythmia (composite); POTs; Dysrhythmia | Heart Rate Abnormalities |
| | Acute Coronary Syndrome; Unstable angina; Acute MI; Acute myocardial infarction; myocardial infarction | Acute Coronary Disease |
| Neurological/neuromuscular/ Nervous system functioning, symptoms and conditions (4) | Stroke; Cerebrovascular disorders; Cerebrovascular disorders (composite); Acute ischemic or embolic stroke; Stroke (composite); Stroke and Sequelae (composite) | Cerebrovascular disorders (unspecified) |
| | Haemorrhagic stroke; Hemorrhagic stroke; Intracerebral hemorrhage | Hemorrhagic stroke |
| | Ischemic stroke; Ischaemic stroke; Stroke and Sequelae (composite) | Ischemic stroke |
| | Movement disorders; Extrapyramidal/ movement disorders; | Movement disorders |
| | Peripheral neuropathy; Peripheral Nerve Conditions; Peripheral neuropathies (composite); Peripheral Nerve Disorders | Peripheral neuropathy |
| | Epilepsy and seizures, Seizure, Epilepsy, Epilepsy; convulsions | Seizure/epilepsy |
| | Myoneural Conditions; Myoclonus; Myasthenia gravis | Myoneural Disorders |
| Psychiatric, Mental functioning, symptoms and conditions (6) | Anxiety/depression; Anxiety; Anxiety disorders; Depression, Major depressive disorders | Anxiety/depression |
| Respiratory, Respiratory functioning, symptoms and conditions (7) | Pulmonary embolism, Acute pulmonary embolism | Pulmonary embolism, Acute pulmonary embolism |

| | | |
|---|---|---|
| | Acute Respiratory Failure, ARDS; Chronic Respiratory Failure; Respiratory Failure (composite), Respiratory insufficiency; | Respiratory Failure |
| Metabolic, Other (13) | Type 2 Diabetes | Diabetes mellitus type 2 (DM Type 2) |

**Note: The numbers in brackets correspond to the codes explained in Tables S5.**

**Table S8. Post-acute outcomes included in the meta-analysis, by study.**

| Author | COVID-19 group (n) | ILI group (n) | Frequency of outcome (COVID-19 group) | Frequency of outcome (ILI group) | Outcome | Symptom(1)/ Condition(2) | Consensus code* |
|---|---|---|---|---|---|---|---|
| Baskett W et al. December, 2022. Columbia, USA | 17487 | 17487 | 2699 | 2382 | Abnormal breathing | 1 | 7 |
| Donnachie E et al. September 2022. Munchen, Germany | 348087 | 55641 | 37237 | 3452 | Abnormal breathing | 1 | 7 |
| Fung KW et al. April, 2023. Bethesda, USA | 293172 | 140697 | 91477 | 34875 | Abnormal breathing | 1 | 7 |
| Liu TH et al. November, 2023. Tainan, Taiwan | 9220 | 9220 | 660 | 328 | Abnormal breathing | 1 | 7 |
| Liu, Ting-Hui et al. December, 2023. Tainan, Taiwan | 6614 | 6614 | 423 | 282 | Abnormal breathing | 1 | 7 |
| Tesch F et al. October, 2024. Dresden, Germany | 547465 | 518706 | 11880 | 4409 | Abnormal breathing | 1 | 7 |
| Velásquez EE et al. July, 2024. Palo Alto, USA | 26732 | 26826 | 1217 | 889 | Abnormal breathing | 1 | 7 |
| Baskett W et al. December, 2022. Columbia, USA | 17487 | 17487 | 608 | 574 | Acute Coronary Syndrome | 2 | 1 |
| Daugherty S E et al. April, 2021. Minneapollis, USA | 181613 | 181613 | 363 | 381 | Acute Coronary Syndrome | 2 | 1 |
| Ward A et al. January, 2022. California, USA | 417153 | 344205 | 4094 | 2008 | Acute Coronary Syndrome | 2 | 1 |

| | | | | | | | |
|---|---|---|---|---|---|---|---|
| Wee E L et al. April, 2025. Singapore, Singapore | 69649 | 11871 | 946 | 163 | Acute Coronary Syndrome | 2 | 1 |
| Baskett W et al. December, 2022. Columbia, USA | 17487 | 17487 | 5547 | 5553 | Anxiety/depression | 2 | 6 |
| Daugherty S E et al. April, 2021. Minneapollis, USA | 181613 | 181613 | 7891 | 7465 | Anxiety/depression | 2 | 6 |
| Liu TH et al. November, 2023. Tainan, Taiwan | 9220 | 9220 | 360 | 285 | Anxiety/depression | 2 | 6 |
| Liu, Ting-Hui et al. December, 2023. Tainan, Taiwan | 6614 | 6614 | 487 | 437 | Anxiety/depression | 2 | 6 |
| Wee L E et al. Dec, 2023. Singapore, Singapore | 477086 | 1272501 | 971 | 3025 | Anxiety/depression | 2 | 6 |
| Havenon A et al. April,2024. New Haven, USA | 77272 | 77272 | 1522 | 1857 | Cerebrovascular disorders (unespecified) | 2 | 4 |
| Velásquez EE et al. July, 2024. Palo Alto, USA | 27825 | 27838 | 80 | 76 | Cerebrovascular disorders (unespecified) | 2 | 4 |
| Ward A et al. January, 2022. California, USA | 417153 | 344205 | 3486 | 1217 | Cerebrovascular disorders (unespecified) | 2 | 4 |
| Wee E L et al. April, 2025. Singapore, Singapore | 70435 | 11985 | 630 | 88 | Cerebrovascular disorders (unespecified) | 2 | 4 |

| | | | | | | | |
|---|---|---|---|---|---|---|---|
| Wee L E et al. Dec, 2023. Singapore, Singapore | 474538 | 1275525 | 1084 | 2394 | Cerebrovascular disorders (unespecified) | 2 | 4 |
| Baskett W et al. December, 2022. Columbia, USA | 17487 | 17487 | 2395 | 2227 | Chest/throat pain | 1 | 7 |
| Fung KW et al. April, 2023. Bethesda, USA | 293172 | 140697 | 51470 | 25603 | Chest/throat pain | 1 | 7 |
| Liu TH et al. November, 2023. Tainan, Taiwan | 9220 | 9220 | 274 | 222 | Chest/throat pain | 1 | 7 |
| Liu, Ting-Hui et al. December, 2023. Tainan, Taiwan | 6614 | 6614 | 196 | 161 | Chest/throat pain | 1 | 7 |
| Tesch F et al. October, 2024. Dresden, Germany | 507500 | 530000 | 203 | 53 | Chest/throat pain | 1 | 7 |
| Fung KW et al. April, 2023. Bethesda, USA | 293172 | 140697 | 57201 | 36439 | Cough | 1 | 7 |
| Liu TH et al. November, 2023. Tainan, Taiwan | 9220 | 9220 | 271 | 219 | Cough | 1 | 7 |
| Liu, Ting-Hui et al. December, 2023. Tainan, Taiwan | 6614 | 6614 | 134 | 124 | Cough | 1 | 7 |
| Baskett W et al. December, 2022. Columbia, USA | 17487 | 17487 | 2321 | 2586 | Cough | 1 | 7 |
| Baskett W et al. December, 2022. Columbia, USA | 17487 | 17487 | 3826 | 3608 | Diabetes mellitus type 2 (DM Type 2) | 2 | 13 |

| | | | | | | | |
|---|---|---|---|---|---|---|---|
| Daugherty S E et al. April, 2021. Minneapollis, USA | 181613 | 181613 | 1906 | 1398 | Diabetes mellitus type 2 (DM Type 2) | 2 | 13 |
| Velásquez EE et al. July, 2024. Palo Alto, USA | 23934 | 23978 | 2954 | 2779 | Diabetes mellitus type 2 (DM Type 2) | 2 | 13 |
| Wee E L et al. April, 2025. Singapore, Singapore | 67136 | 11509 | 1319 | 237 | Diabetes mellitus type 2 (DM Type 2) | 2 | 13 |
| Baskett W et al. December, 2022. Columbia, USA | 17487 | 17487 | 2184 | 2006 | Fatigue/malaise | 1 | 2 |
| Daugherty S E et al. April, 2021. Minneapollis, USA | 181613 | 181613 | 8790 | 6592 | Fatigue/malaise | 1 | 2 |
| Donnachie E et al. September 2022. Munchen, Germany | 348087 | 55641 | 49126 | 6051 | Fatigue/malaise | 1 | 2 |
| Fung KW et al. April, 2023. Bethesda, USA | 293172 | 140697 | 97381 | 41971 | Fatigue/malaise | 1 | 2 |
| Liu TH et al. November, 2023. Tainan, Taiwan | 9220 | 9220 | 424 | 270 | Fatigue/malaise | 1 | 2 |
| Liu, Ting-Hui et al. December, 2023. Tainan, Taiwan | 6614 | 6614 | 248 | 165 | Fatigue/malaise | 1 | 2 |
| Tesch F et al. October, 2024. Dresden, Germany | 571622 | 540000 | 13670 | 6975 | Fatigue/malaise | 1 | 2 |
| Velásquez EE et al. July, 2024. Palo Alto, USA | 26557 | 26558 | 1043 | 883 | Fatigue/malaise | 1 | 2 |

| | | | | | | | |
|---|---|---|---|---|---|---|---|
| Wee E L et al. April, 2025. Singapore, Singapore | 70534 | 11907 | 500 | 43 | Fatigue/malaise | 1 | 2 |
| Wee L E et al. Dec, 2023. Singapore, Singapore | 479336 | 1281784 | 371 | 637 | Fatigue/malaise | 1 | 2 |
| Baskett W et al. December, 2022. Columbia, USA | 17487 | 17487 | 2557 | 2583 | Headache/migraine | 1 | 4 |
| Daugherty S E et al. April, 2021. Minneapollis, USA | 181613 | 181613 | 1398 | 1416 | Headache/migraine | 1 | 4 |
| Fung KW et al. April, 2023. Bethesda, USA | 293172 | 140697 | 20976 | 11925 | Headache/migraine | 1 | 4 |
| Havenon A et al. April,2024. New Haven, USA | 77272 | 77272 | 1519 | 3957 | Headache/migraine | 1 | 4 |
| Liu TH et al. November, 2023. Tainan, Taiwan | 9220 | 9220 | 137 | 128 | Headache/migraine | 1 | 4 |
| Liu, Ting-Hui et al. December, 2023. Tainan, Taiwan | 6614 | 6614 | 74 | 72 | Headache/migraine | 1 | 4 |
| Velásquez EE et al. July, 2024. Palo Alto, USA | 27022 | 27075 | 650 | 487 | Headache/migraine | 1 | 4 |
| Wee E L et al. April, 2025. Singapore, Singapore | 70577 | 11993 | 167 | 22 | Headache/migraine | 1 | 4 |
| Wee L E et al. Dec, 2023. Singapore, Singapore | 479338 | 1283121 | 350 | 908 | Headache/migraine | 1 | 4 |
| Daugherty S E et al. April, 2021. Minneapollis, USA | 181613 | 181613 | 2761 | 1979 | Heart Rate Abnormalities | 1 | 1 |
| Baskett W et al. December, 2022. Columbia, USA | 17487 | 17487 | 2158 | 1949 | Heart Rate Abnormalities | 2 | 1 |

| | | | | | | | |
|---|---|---|---|---|---|---|---|
| Velásquez EE et al. July, 2024. Palo Alto, USA | 27413 | 27488 | 437 | 316 | Heart Rate Abnormalities | 2 | 1 |
| Wee E L et al. April, 2025. Singapore, Singapore | 69934 | 11903 | 923 | 144 | Heart Rate Abnormalities | 2 | 1 |
| Daugherty S E et al. April, 2021. Minneapollis, USA | 181613 | 181613 | 109 | 91 | Hemorrhagic stroke | 2 | 4 |
| Baskett W et al. December, 2022. Columbia, USA | 17487 | 17487 | 26 | 30 | Hemorrhagic stroke | 2 | 4 |
| Wee L E et al. Dec, 2023. Singapore, Singapore | 478770 | 1283790 | 322 | 689 | Hemorrhagic stroke | 2 | 4 |
| Zarifkar P et al. June,2022. Copenhagen, Denmark | 43262 | 8102 | 16 | 1 | Hemorrhagic stroke | 2 | 4 |
| Baskett W et al. December, 2022. Columbia, USA | 17487 | 17487 | 467 | 435 | Ischemic stroke | 2 | 4 |
| Daugherty S E et al. April, 2021. Minneapollis, USA | 181613 | 181613 | 345 | 272 | Ischemic stroke | 2 | 4 |
| Wee L E et al. Dec, 2023. Singapore, Singapore | 476972 | 1280440 | 625 | 1370 | Ischemic stroke | 2 | 4 |
| Zarifkar P et al. June,2022. Copenhagen, Denmark | 43262 | 8102 | 279 | 65 | Ischemic stroke | 2 | 4 |
| Baskett W et al. December, 2022. Columbia, USA | 17487 | 17487 | 52 | 60 | Loss of taste/smell | 1 | 4 |
| Cabrera C I et al. November, 2023. Cleveland, USA | 2544070 | 101886 | 1108 | 180 | Loss of taste/smell | 1 | 4 |
| Daugherty S E et al. April, 2021. Minneapollis, USA | 181613 | 181613 | 399 | 73 | Loss of taste/smell | 1 | 4 |

| | | | | | | | |
|---|---|---|---|---|---|---|---|
| Donnachie E et al. September 2022. Munchen, Germany | 348087 | 55641 | 11435 | 725 | Loss of taste/smell | 1 | 4 |
| Fung KW et al. April, 2023. Bethesda, USA | 293172 | 140697 | 1825 | 456 | Loss of taste/smell | 1 | 4 |
| Liu TH et al. November, 2023. Tainan, Taiwan | 9220 | 9220 | 2 | 3 | Loss of taste/smell | 1 | 4 |
| Liu, Ting-Hui et al. December, 2023. Tainan, Taiwan | 6614 | 6614 | 0 | 0 | Loss of taste/smell | 1 | 4 |
| Wee E L et al. April, 2025. Singapore, Singapore | 70281 | 11983 | 332 | 45 | Loss of taste/smell | 1 | 4 |
| Wee L E et al. Dec, 2023. Singapore, Singapore | 475499 | 1275619 | 995 | 2357 | Loss of taste/smell | 1 | 4 |
| Baskett W et al. December, 2022. Columbia, USA | 17487 | 17487 | 1299 | 1160 | Memory loss/brain fog | 1 | 5 |
| Daugherty S E et al. April, 2021. Minneapollis, USA | 181613 | 181613 | 1307 | 872 | Memory loss/brain fog | 1 | 5 |
| Donnachie E et al. September 2022. Munchen, Germany | 348087 | 55641 | 1507 | 149 | Memory loss/brain fog | 1 | 5 |
| Fung KW et al. April, 2023. Bethesda, USA | 293172 | 140697 | 17874 | 7817 | Memory loss/brain fog | 1 | 5 |
| Liu TH et al. November, 2023. Tainan, Taiwan | 9220 | 9220 | 224 | 136 | Memory loss/brain fog | 1 | 5 |
| Liu, Ting-Hui et al. December, 2023. Tainan, Taiwan | 6614 | 6614 | 115 | 63 | Memory loss/brain fog | 1 | 5 |
| Tesch F et al. October, 2024. Dresden, Germany | 565870 | 543077 | 2603 | 2118 | Memory loss/brain fog | 1 | 5 |

| | | | | | | | |
|---|---|---|---|---|---|---|---|
| Velásquez EE et al. July, 2024. Palo Alto, USA | 27002 | 27009 | 590 | 620 | Memory loss/brain fog | 1 | 5 |
| Wee E L et al. April, 2025. Singapore, Singapore | 70127 | 11972 | 1005 | 113 | Memory loss/brain fog | 1 | 5 |
| Wee L E et al. Dec, 2023. Singapore, Singapore | 476382 | 1280986 | 1710 | 3076 | Memory loss/brain fog | 1 | 5 |
| Havenon A et al. April,2024. New Haven, USA | 77272 | 77272 | 1132 | 1900 | Movement disorders | 2 | 4 |
| Wee E L et al. April, 2025. Singapore, Singapore | 70520 | 11993 | 178 | 15 | Movement disorders | 2 | 4 |
| Wee L E et al. Dec, 2023. Singapore, Singapore | 478520 | 1282805 | 280 | 665 | Movement disorders | 2 | 4 |
| Baskett W et al. December, 2022. Columbia, USA | 17487 | 17487 | 4085 | 3782 | Myalgia/arthralgia | 1 | 3 |
| Daugherty S E et al. April, 2021. Minneapollis, USA | 181613 | 181613 | 8227 | 7682 | Myalgia/arthralgia | 1 | 3 |
| Donnachie E et al. September 2022. Munchen, Germany | 348087 | 55641 | 14517 | 2335 | Myalgia/arthralgia | 1 | 3 |
| Fung KW et al. April, 2023. Bethesda, USA | 293172 | 140697 | 16200 | 9047 | Myalgia/arthralgia | 1 | 3 |
| Liu TH et al. November, 2023. Tainan, Taiwan | 9220 | 9220 | 57 | 69 | Myalgia/arthralgia | 1 | 3 |
| Liu, Ting-Hui et al. December, 2023. Tainan, Taiwan | 6614 | 6614 | 33 | 24 | Myalgia/arthralgia | 1 | 3 |
| Velásquez EE et al. July, 2024. Palo Alto, USA | 27846 | 27886 | 66 | 52 | Myoneural Disorders | 2 | 4 |

| | | | | | | | |
|---|---|---|---|---|---|---|---|
| Wee L E et al. Dec, 2023. Singapore, Singapore | 480022 | 1281854 | 15 | 19 | Myoneural Disorders | 2 | 4 |
| Zarifkar P et al. June,2022. Copenhagen, Denmark | 43262 | 8102 | 1 | 0 | Myoneural Disorders | 2 | 4 |
| Baskett W et al. December, 2022. Columbia, USA | 17487 | 17487 | 806 | 594 | Palpitations | 1 | 1 |
| Fung KW et al. April, 2023. Bethesda, USA | 293172 | 140697 | 39587 | 15994 | Palpitations | 1 | 1 |
| Liu TH et al. November, 2023. Tainan, Taiwan | 9220 | 9220 | 90 | 83 | Palpitations | 1 | 1 |
| Liu, Ting-Hui et al. December, 2023. Tainan, Taiwan | 6614 | 6614 | 43 | 37 | Palpitations | 1 | 1 |
| Baskett W et al. December, 2022. Columbia, USA | 17487 | 17487 | 669 | 616 | Peripheral neuropathy | 2 | 4 |
| Daugherty S E et al. April, 2021. Minneapollis, USA | 181613 | 181613 | 581 | 363 | Peripheral neuropathy | 2 | 4 |
| Havenon A et al. April,2024. New Haven, USA | 77272 | 77272 | 1460 | 2793 | Peripheral neuropathy | 2 | 4 |
| Velásquez EE et al. July, 2024. Palo Alto, USA | 27540 | 27580 | 254 | 254 | Peripheral neuropathy | 2 | 4 |
| Wee E L et al. April, 2025. Singapore, Singapore | 70554 | 11999 | 152 | 34 | Peripheral neuropathy | 2 | 4 |
| Wee L E et al. Dec, 2023. Singapore, Singapore | 478007 | 1283132 | 402 | 1006 | Peripheral neuropathy | 2 | 4 |
| Baskett W et al. December, 2022. Columbia, USA | 17487 | 17487 | 201 | 165 | Pulmonary embolism | 2 | 7 |

| | | | | | | | |
|---|---|---|---|---|---|---|---|
| Daugherty S E et al. April, 2021. Minneapollis, USA | 181613 | 181613 | 363 | 236 | Pulmonary embolism | 2 | 7 |
| Donnachie E et al. September 2022. Munchen, Germany | 348087 | 55641 | 1620 | 178 | Pulmonary embolism | 2 | 7 |
| Tesch F et al. October, 2024. Dresden, Germany | 558182 | 538889 | 1228 | 485 | Pulmonary embolism | 2 | 7 |
| Ward A et al. January, 2022. California, USA | 417153 | 344205 | 2315 | 772 | Pulmonary embolism | 2 | 7 |
| Baskett W et al. December, 2022. Columbia, USA | 17487 | 17487 | 903 | 781 | Respiratory Failure | 2 | 7 |
| Daugherty S E et al. April, 2021. Minneapollis, USA | 181613 | 181613 | 418 | 490 | Respiratory Failure | 1 | 7 |
| Tesch F et al. October, 2024. Dresden, Germany | 565762 | 547714 | 3338 | 1917 | Respiratory Failure | 2 | 7 |
| Daugherty S E et al. April, 2021. Minneapollis, USA | 181613 | 181613 | 254 | 254 | Seizure/epilepsy | 1 | 4 |
| Havenon A et al. April,2024. New Haven, USA | 77272 | 77272 | 1218 | 1649 | Seizure/epilepsy | 2 | 4 |
| Velásquez EE et al. July, 2024. Palo Alto, USA | 27867 | 27889 | 56 | 46 | Seizure/epilepsy | 2 | 4 |
| Wee L E et al. Dec, 2023. Singapore, Singapore | 478337 | 1283112 | 152 | 325 | Seizure/epilepsy | 2 | 4 |
| Baskett W et al. December, 2022. Columbia, USA | 17487 | 17487 | 1168 | 1186 | Sleep disorders | 1 | 6 |
| Fung KW et al. April, 2023. Bethesda, USA | 293172 | 140697 | 2868 | 1285 | Sleep disorders | 1 | 6 |

| | | | | | | | |
|---|---|---|---|---|---|---|---|
| Liu TH et al. November, 2023. Tainan, Taiwan | 9220 | 9220 | 92 | 69 | Sleep disorders | 1 | 6 |
| Liu, Ting-Hui et al. December, 2023. Tainan, Taiwan | 6614 | 6614 | 82 | 68 | Sleep disorders | 1 | 6 |
| Velásquez EE et al. July, 2024. Palo Alto, USA | 26491 | 26524 | 1000 | 882 | Sleep disorders | 1 | 6 |
| Wee L E et al. Dec, 2023. Singapore, Singapore | 476037 | 1274090 | 574 | 1650 | Sleep disorders | 1 | 6 |
| Baskett W et al. December, 2022. Columbia, USA | 17487 | 17487 | 156 | 130 | Visual and Auditory Disturbance | 1 | 4 |
| Velásquez EE et al. July, 2024. Palo Alto, USA | 27810 | 27818 | 87 | 100 | Visual and Auditory Disturbance | 1 | 4 |
| Wee L E et al. Dec, 2023. Singapore, Singapore | 477051 | 1278300 | 954 | 2311 | Visual and Auditory Disturbance | 1 | 4 |

***Note: Consensus codes from Tables S5.**

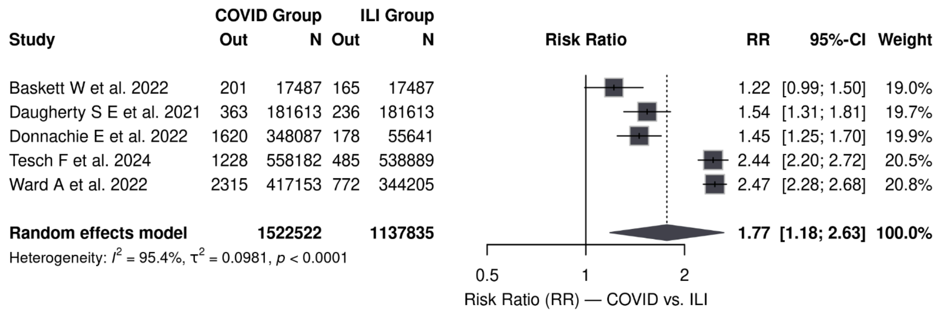


**Figure S1. Pooled risk of pulmonary embolism (COVID vs ILI).** Forest plot of five studies comparing the risk of persistent respiratory symptoms after SARS-CoV-2 infection versus other respiratory viral infections. Boxes = study point estimates; horizontal lines = 95% CIs; diamond = pooled RR (random effects). Pooled RR 1.7 (95% CI 1.18–2.63); I² = 95.4%.

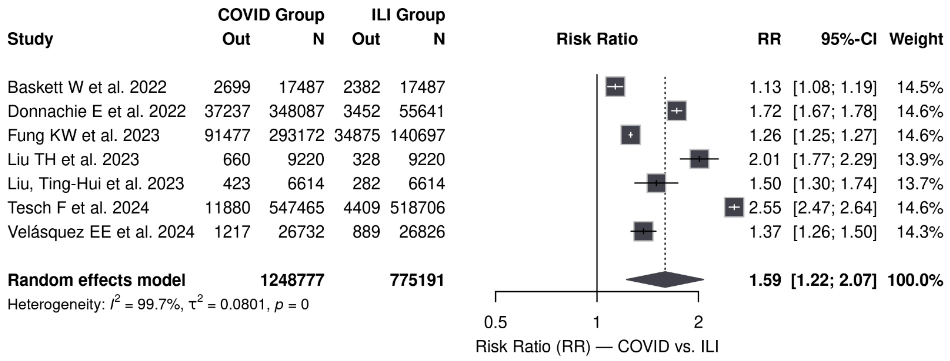


**Figure S2. Pooled risk of abnormal breathing (COVID vs ILI).** Forest plot of seven studies comparing the risk of persistent respiratory symptoms after SARS-CoV-2 infection versus other respiratory viral infections. Boxes = study point estimates; horizontal lines = 95% CIs; diamond = pooled RR (random effects). Pooled RR 1.59 (95% CI 1.22–2.07); I² = 99.7%.

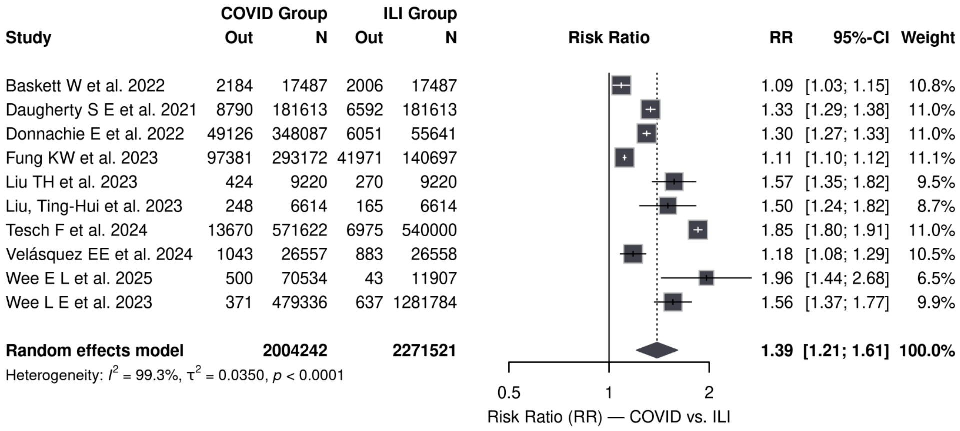


**Figure S3. Pooled risk of fatigue/malaise (COVID vs ILI).** Forest plot of ten studies comparing the risk of persistent respiratory symptoms after SARS-CoV-2 infection versus other respiratory viral infections. Boxes = study point estimates; horizontal lines = 95% CIs; diamond = pooled RR (random effects). Pooled RR 1.59 (95% CI 1.22–1.61); I² = 99.3%.

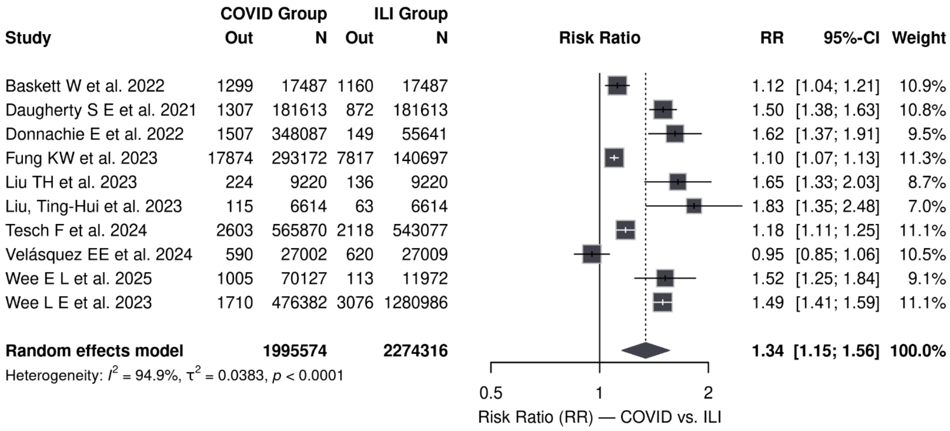


**Figure S4. Pooled risk of memory loss/brain fog (COVID vs ILI).** Forest plot of ten studies comparing the risk of persistent respiratory symptoms after SARS-CoV-2 infection versus other respiratory viral infections. Boxes = study point estimates; horizontal lines = 95% CIs; diamond = pooled RR (random effects). Pooled RR 1.34 (95% CI 1.15–1.56); I² = 94.9%.

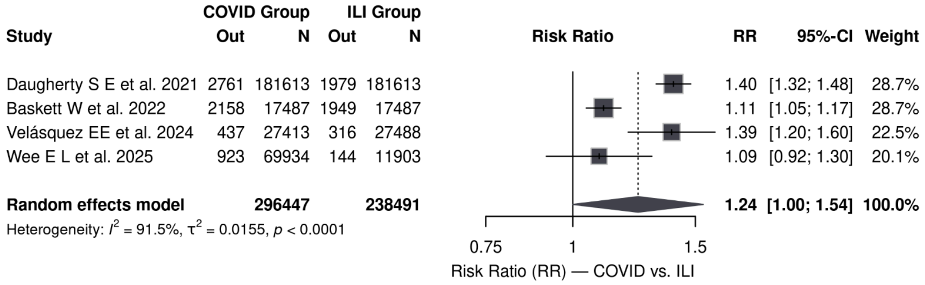


**Figure S5. Pooled risk of heart rate abnormalities (COVID vs ILI).** Forest plot of four studies comparing the risk of persistent respiratory symptoms after SARS-CoV-2 infection versus other respiratory viral infections. Boxes = study point estimates; horizontal lines = 95% CIs; diamond = pooled RR (random effects). Pooled RR 1.24 (95% CI 1.00–1.54); I² = 91.5%.

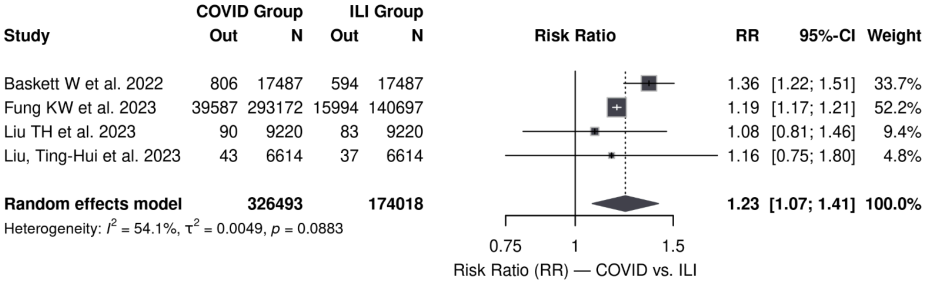


**Figure S6. Pooled risk of palpitations (COVID vs ILI).** Forest plot of four studies comparing the risk of persistent respiratory symptoms after SARS-CoV-2 infection versus other respiratory viral infections. Boxes = study point estimates; horizontal lines = 95% CIs; diamond = pooled RR (random effects). Pooled RR 1.23 (95% CI 1.07–1.41); I² = 54.1%.

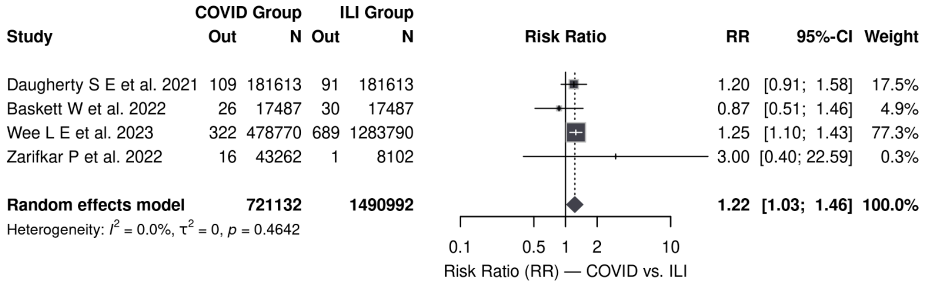


**Figure S7. Pooled risk of hemorrhagic stroke (COVID vs ILI).** Forest plot of four studies comparing the risk of persistent respiratory symptoms after SARS-CoV-2 infection versus other respiratory viral infections. Boxes = study point estimates; horizontal lines = 95% CIs; diamond = pooled RR (random effects). Pooled RR 1.22 (95% CI 1.03–1.46); I² = 0.0%.

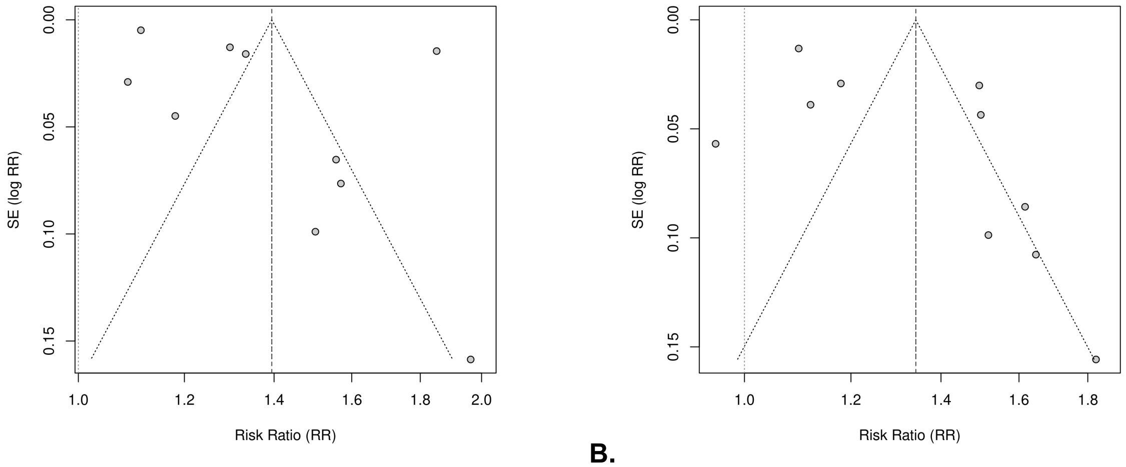


**B.**

**Figure S8. Funnel plots for outcomes with ≥10 studies.** Funnel plots showing the relationship between risk ratios (RRs) and the standard error of the log risk ratio (SE[log RR]) for studies assessing fatigue/malaise (**A**) and memory loss/brain fog (**B**) following COVID-19 compared with influenza-like illness (ILI). Formal tests for funnel plot asymmetry (Egger’s and Begg's effect estimate, and diagonal lines indicate pseudo 95% confidence limits. Both plots show at most minor visual asymmetry. Formal tests for funnel plot asymmetry (Egger’s and Begg’s tests) did not indicate statistically significant small-study effects.

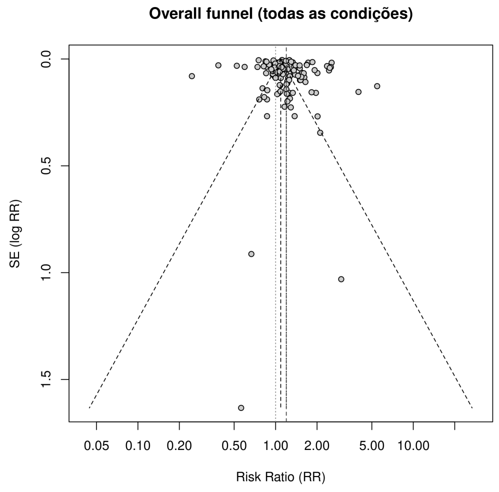


**Figure S9. Overall funnel plot including all conditions.** Funnel plot of risk ratios (RRs) plotted against SE(log RR) for all clinical conditions included in the meta-analysis. The vertical line indicates the null value (RR = 1). Owing to the clinical and methodological heterogeneity across outcomes, this plot is presented for descriptive purposes only.

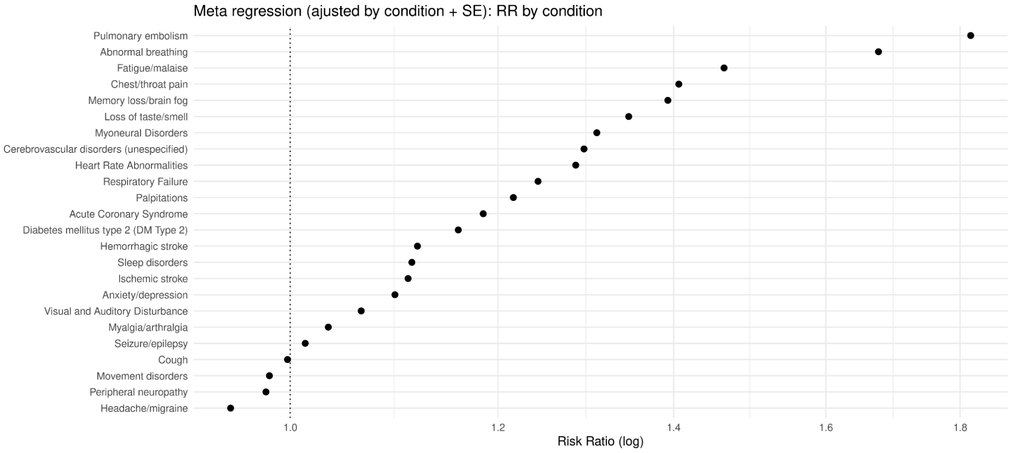


**Figure S10. Meta-regression coefficient plot adjusted for study precision.** Coefficient plot from the meta-regression model including clinical condition and standardized SE(log RR) as moderators. Points represent adjusted risk ratios (RRs) for each condition on the logarithmic scale, relative to the null value (RR = 1). The persistence of differences between conditions after adjustment suggests that variability across outcomes is not driven by study precision.

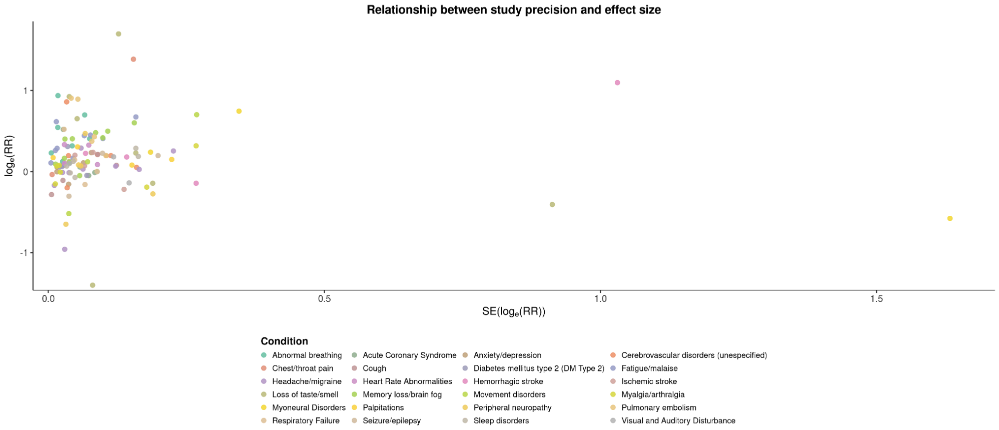


**Figure S11. Relationship between study precision and effect size.** Scatter and bubble plots illustrating the relationship between the standard error of the log risk ratio (SE[log RR]) and log RR across all included studies, stratified by clinical condition. The absence of a clear directional trend is consistent with the meta-regression results and suggests no systematic association between study precision and effect magnitude.

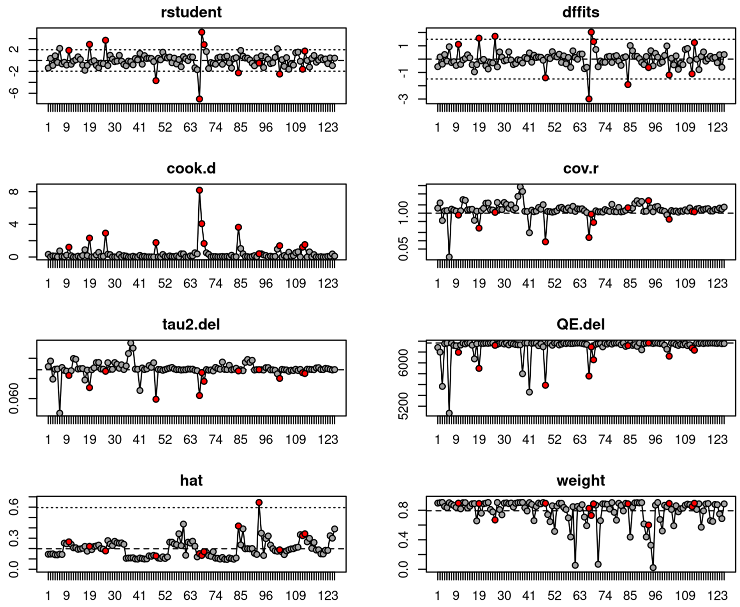


**Figure S12. Influence diagnostics and sensitivity analyses.** Influence diagnostics from the random-effects meta-analysis, including standardized residuals (rstudent), Cook's distance, DFFITS, covariance ratios, changes in between-study variance ($\tau^2$), and Cochran's Q statistic following study deletion. Highlighted points indicate observations exceeding conventional thresholds. Overall, no individual study exerted disproportionate influence on the pooled estimates, supporting the robustness of the results.
